# Lineage-specific protection and immune imprinting shape the age distributions of influenza B cases

**DOI:** 10.1101/2020.09.30.20204909

**Authors:** Marcos C. Vieira, Celeste M. Donato, Philip Arevalo, Guus F. Rimmelzwaan, Timothy Wood, Liza Lopez, Q. Sue Huang, Vijaykrishna Dhanasekaran, Katia Koelle, Sarah Cobey

## Abstract

How a history of influenza virus infections contributes to protection is not fully understood, but such protection might explain the contrasting age distributions of cases of the two lineages of influenza B, B/Victoria and B/Yamagata. Fitting a statistical model to those distributions using surveillance data from New Zealand, we found they could be explained by historical changes in lineage frequencies combined with cross-protection between strains of the same lineage. We found additional protection against B/Yamagata in people for whom it was their first influenza B infection, similar to the immune imprinting observed in influenza A. While the data were not informative about B/Victoria imprinting, B/Yamagata imprinting could explain the fewer B/Yamagata than B/Victoria cases in cohorts born in the 1990s and the bimodal age distribution of B/Yamagata cases. Longitudinal studies can test if these forms of protection inferred from historical data extend to more recent strains and other populations.

## Introduction

The incidence of pathogens that induce long-lasting immunity, such as measles or mumps, typically peaks at a young age and decreases as hosts gain immune protection with age (Galbraith et al., 1984; Anderson et al., 1987; Grenfell and Anderson, 1985). However, many pathogens can infect the same host multiple times due to the circulation of antigenically distinct strains in the population. Changes in the prevalences of strains over time can lead to different infection histories in hosts born at different times, and such differences in infection history can lead to complicated age distributions of infection (Gupta et al., 1998; Kucharski and Gog, 2012a,b). Influenza viruses, for instance, circulate as antigenically distinct variants, including the types A and B, subtypes of influenza A, lineages of influenza B, and clades within them. Changes in variant prevalence over time generate different infection histories that are correlated across birth cohorts (Kilbourne, 2006; Rota et al., 1990; Shaw et al., 2002), but how differences in infection history affect protection and shape the age distribution of influenza virus infections is not fully understood.

Early childhood infections with influenza A have long-lasting consequences for protection against influenza A subtypes, a phenomenon termed “immune imprinting” (Gostic et al., 2016, 2019; Arevalo et al., 2020b). Subtypes are distinguished by their surface proteins, hemagglutinin (HA) and neuraminidase (NA), with HA subtypes clustered into the major phylogenetic groups 1 (including, among others, H1, H2 and H5) and 2 (including H3 and H7). Different subtypes of influenza A have circulated in the 20th century (Kilbourne, 2006), and protection against severe infection and death is higher against subtypes whose HAs are genetically similar to the HA of the subtype with which a person was likely first infected (Gostic et al., 2016, 2019; Arevalo et al., 2020b). For instance, early infection with H1N1 or H2N2 is associated with lifelong protection against severe infections with not only H1N1 but also H5N1, and early H3N2 infection protects against severe infection with H7N9 as well as H3N2. Subtypes H1N1 and H2N2 were the only subtypes circulating between 1918 and 1968, and H3N2 has been more common than H1N1 since 1968. Thus, the age distributions of clinical infections with H1N1 and H5N1 skew young, and the distributions of H7N9 and H3N2 infections skew old, because of the lasting impact of childhood immunity to the first HA encountered (Gostic et al., 2016, 2019; Arevalo et al., 2020b).

Despite its durability, imprinting protection does not completely prevent re-infection with the same subtype, and models based on imprinting protection alone cannot completely recapitulate the age distribution of cases (Gostic et al., 2019; Arevalo et al., 2020b). Longitudinal analyses of antibody titers, which reveal subclinical infections, suggest that protection after infection with influenza type A decreases significantly within 3.5-7 years (Kucharski et al., 2015; Ranjeva et al., 2019). Repeated clinical infections of the same subtype have been observed in the same person (Smith and Davies, 1976; Frank and Taber, 1983; Davies et al., 1984; Horby et al., 2012) and are likely enabled by antigenic evolution of HA and NA, which experience strong positive selection (Bush et al., 1999; Ferguson et al., 2003; Smith et al., 2004; Bedford et al., 2014). Thus, protection against infection with a subtype appears to depend not only on imprinting protection (early infection with a subtype) but also cross-protection from recent infections with that subtype. This cross-protection can be sensitive to the precise strains with which a person was infected, apparent as birth cohort effects and reproducible in animal models (Linderman et al., 2014; Huang et al., 2015; Kucharski et al., 2015; Petrie et al., 2016).

Like the different subtypes of influenza A, the two lineages of influenza type B have distinct age distributions of medically attended infections. The B/Victoria and B/Yamagata lineages diverged in the early 1980s (Rota et al., 1990; Dudas et al., 2015) and have circulated with varying frequencies since, causing 25% of global influenza cases detected in 2000-2018 (Caini et al., 2019). While the incidence of medically attended infections is highest in children for both lineages, B/Yamagata appears less common than B/Victoria in teenagers and young adults but the pattern reverses in older age groups. This pattern has been observed in surveillance from the 2000s and 2010s in Oceania (Vijaykrishna et al., 2015), East Asia (Tan et al., 2013), Europe (Sočan et al., 2014; Puzelli et al., 2019) and North America (Skowronski et al., 2017). It has also been observed in isolates from sequence databases (Virk et al., 2020). Changes with age in the expression of sialic acid receptors have been proposed to explain the lower mean age of B/Victoria cases (Vijaykrishna et al., 2015), but this explanation does not account for the higher frequency of B/Yamagata cases in middle-aged people.

Alternatively or in addition to physiological changes, differences in cohorts’ susceptibility to influenza B lineages might arise from differences in cohorts’ infection histories. It is unclear if people have increased protection against their first infecting influenza B lineage. One hypothesized mechanism for immune imprinting in influenza A is antibodies to conserved epitopes (Gostic et al., 2016, 2019; Arevalo et al., 2020a,b; Carreño et al., 2020). Since B/Victoria and B/Yamagata diverged from each other more recently (Rota et al., 1990; Chen and Holmes, 2008; Dudas et al., 2015) and evolve antigenically much more slowly than the influenza A subtypes (Bedford et al., 2014, 2015; Vijaykrishna et al., 2015), conserved epitopes within a lineage might also be conserved between lineages, leading to strong cross-lineage protection by antibodies targeting those epitopes, regardless of which lineage was encountered first. Alternatively, cross-lineage protection may be weak or asymmetrical, with downstream consequences for the age distribution of medically attended cases.

To investigate how protection might arise from infections with B/Victoria and B/Yamagata and contribute to differences in their age distributions, we fitted a statistical model to medically attended influenza B cases recorded by systematic surveillance in New Zealand from 2001 to 2019 (Figure 1). The model used the estimated historical frequencies of the lineages to estimate the probabilities of different infection histories and the strength of within- and cross-lineage protection from previous infections. We found that differences in birth cohorts’ susceptibility to each lineage are consistent with strong cross-protection between strains of the same lineage. In addition to within-lineage protection independent of the order of infection, we found evidence of protection against B/Yamagata in people for whom it was the the first influenza B infection, while the strength of a similar effect for B/Victoria could not be estimated from the data. This imprinting protection against B/Yamagata can explain why cohorts born in the 1990s, when B/Yamagata appears to have been the only lineage circulating in New Zealand, have since had fewer B/Yamagata cases than B/Victoria cases and fewer B/Yamagata cases than older birth cohorts. We found similar support for B/Yamagata imprinting in data from other regions where B/Yamagata was dominant during the 1990s but not in data from China, where B/Victoria was present during that time (Shaw et al., 2002). These results suggest that long-lasting protection from early infections, previously shown for the subtypes and groups of influenza A, also shapes the age distributions of the more recently diverged influenza B lineages.

**Figure 1.**
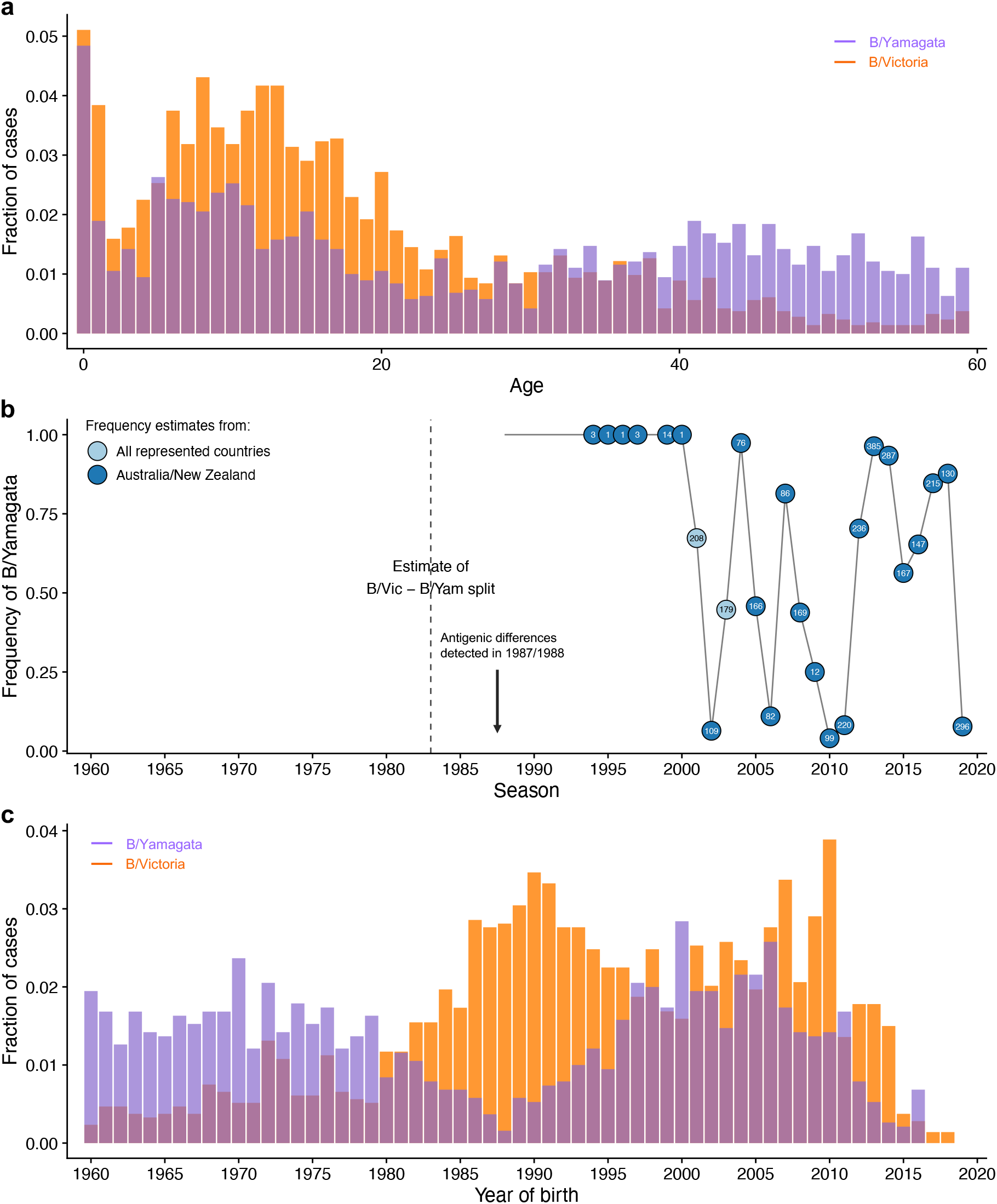
Historical frequencies and age distributions of the influenza B lineages. **a**. Distribution of medically attended influenza B cases in New Zealand in 2001-2019 by age (in years) of the infected person. **b**. Frequency of B/Yamagata estimated from sequences deposited on GISAID and the NCBI Influenza Virus Database. Frequencies were estimated for New Zealand and Australia combined to increase power. The circles are annotated with the number of isolates used to estimate frequencies in each season. In 2001 and 2003, when both lineages were known to be circulating in Australia and New Zealand but the number of isolates from those countries combined were small, we estimated lineage frequencies using isolates from all countries represented in the sequence databases. **c**. Distribution of medically attended influenza B cases in New Zealand in 2001-2019 by birth year of the infected person. In panels **a** and **c**, the fraction of cases was calculated relative to all cases observed for each lineage (including ages and birth years not shown in the figure).

## Results

### Statistical model of influenza B cases by birth year

To test how age, early infections, and protection within and between lineages might shape the distribution of influenza B cases, we fitted a statistical model to medically attended infections detected by influenza surveillance from 2001 to 2019 in New Zealand (Methods: “Case data”; Figure 1, Figure S1). Data from 2002-2013 were previously analyzed by Vijaykrishna et al. (2015). Most of the data (60% of the 4,036 cases identified at the level of the influenza B lineage) were collected by general practices as part of a surveillance program with well-defined sampling criteria independent of the patient’s age (Huang et al., 2008), thus approximating the true age distribution of medically attended infections. The remaining cases were sampled without consistent criteria from hospital samples, mostly inpatient. We performed the main analyses on the entire New Zealand data set but assessed the effect of removing the non-surveillance data and fitting the model only to the general practice surveillance data. In addition to the New Zealand data, we analyzed age distributions from two data sets with unclear sampling criteria: Australian hospital samples compiled by Vijaykrishna et al. (2015) and influenza B isolates from other countries that were submitted to the GISAID database.

Following work on influenza A (Gostic et al., 2016, 2019; Arevalo et al., 2020b), the model describes the probability that a case occurs in each birth cohort (Figure 2; Methods: “Statistical model of influenza B susceptibility based on infection history”). These probabilities are based on:

**Figure 2.**
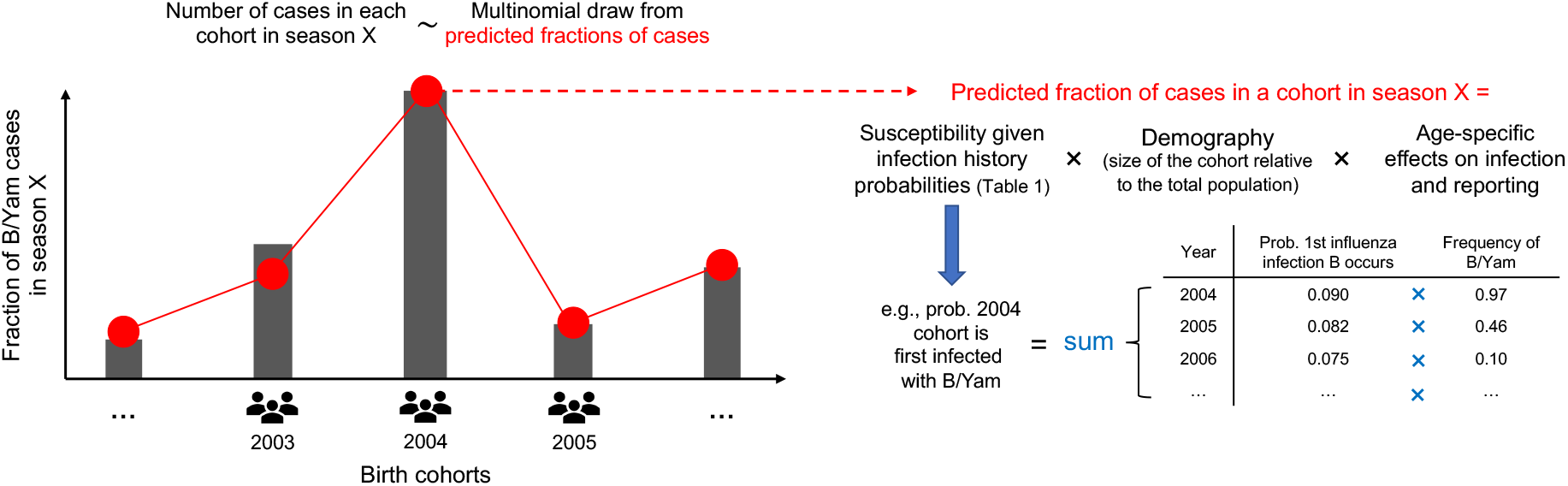
Statistical model of influenza B infections by birth year. Extending the model developed by Gostic et al. (2016), for each influenza B season we predicted the fraction of cases observed in each cohort based on cohort-specific infection histories and on additional factors. The probabilities of the different infection histories in Table 1 are derived in the Methods using a discrete-time probabilistic model of infection. As an example, the diagram shows how the probability that a person is first infected with B/Yamagata depends on the frequency of B/Yamagata in the years after birth. Given a person’s first influenza B infection occurs in a particular year, we assumed that the probability this infection was caused by a particular lineage is equal to the frequency of the lineage in that year. We obtained the total probability of being first infected with a particular lineage by summing across all possible years when the first influenza B infection might have occurred. The susceptibility of each cohort is then calculated as a weighted sum of the susceptibilities associated with each infection history. In addition to infection history, other factors that affect the fraction of cases observed in a birth cohort include the cohort’s size relative to the total population and the effect of age itself (rather than year of birth) on infection risk and on the probability that an infection receives medical attention and becomes a case. We fitted the model by maximum likelihood assuming the distributions of cases by birth year in different seasons were independent multinomial draws

1. *Demography*. The fraction of the total population that belongs to a birth cohort gives a baseline probability that a case occurs in that cohort. We obtained those fractions from demographic data for New Zealand (Methods: “Demographic data”).
2. *Age-specific effects on infection risk*. We assumed that preschoolers (0-5 years old), school-age children and teenagers (6-17 years old) and people 18 years and older had different baseline probabilities of infection, representing differences in exposure and susceptibility related with age itself and not infection history. These baseline probabilities were then modified by protection from previous infections. We compared the estimates for the preschool and school-age infection probabilities with independent estimates.
3. *Age-specific effects on reporting*. We assumed infections in children younger than 2 years old had a different probability of receiving medical attention, and thus being reported as a case, than infections in the rest of the population (estimates of protection were similar when we assumed this effect applied to children younger than 5). We estimated this differential reporting from the case data.
4. *Infection history*. The probability of observing a case of a lineage in a birth cohort could be modified by the average susceptibility of people in that cohort to that lineage, based on the birth cohort’s infection history. Susceptibility is defined as the probability of infection given exposure, relative to that of a naive person. We estimated the probabilities of different infection histories for each birth cohort using a discrete-time model in which the probability of becoming infected with either lineage depends on the lineages’ historical frequencies (Methods: “Infection history probabilities”), which we estimated using sequence databases (Methods: “Historical frequencies of influenza B lineages”). Consistent with historical surveillance and with trends observed in other countries outside East Asia (Shaw et al., 2002), we found that B/Yamagata was the only lineage circulating in New Zealand in the 1990s, although this observation was based on few isolates. B/Victoria started circulating again in the early 2000s, and the two lineages have alternated in dominance since (Figure 1b, Figure S2). We investigated the sensitivity of the results to the frequency of B/Yamagata in the 1990s by alternatively estimating lineage frequencies from all countries represented in sequence databases for that period. We examined four effects of past infection (Table 1):
  a. *Within-lineage protection*. Any previous infection with B/Victoria or B/Yamagata decreases susceptibility to future infections with the same lineage by fractions *χ*_*VV*_ and *χ*_*YY*_, respectively, with a value of 1 representing full protection and a value of 0 representing no protection.
  b. *Cross-lineage protection* Cross-lineage protection was estimated as a fraction (*γ*) of the corresponding within-lineage protection against that lineage: *χ*_*YV*_ = *γ*_*YV*_ × *χ*_*YY*_ and *χ*_*VY*_ = *γ*_*VY*_ ×*χ*_*VV*_, where *χ*_*YV*_ is the protection from B/Victoria against B/Yamagata and *χ*_*VY*_ is the protection from B/Yamagata against B/Victoria. For instance, *γ*_*YV*_ = 0.7 means that compared with within-lineage B/Yamagata protection, protection from B/Victoria against B/Yamagata is 70% as strong. We assumed that once a person was infected with a lineage, within-lineage protection superseded protection from previous infection with the other lineage.
  c. *Lineage-specific imprinting*. To represent increased protection against the lineage of first infection, susceptibility to B/Victoria and B/Yamagata could be further reduced by *R*_*V*_ or *R*_*Y*_ if a person was first infected with the corresponding lineage. Thus, susceptibility to B/Victoria in people first infected with B/Victoria was (1−*χ*_*VV*_)(1− *R*_*V*_), and susceptibility to B/Yamagata in people first infected with B/Yamagata was (1 −*χ*_*YY*_)(1 − *R*_*Y*_). A value of 1 for *R*_*V*_ or *R*_*Y*_ results in perfect protection against the lineage first encountered, whereas a value of 0 means that within-lineage protection is the same regardless of the order of infection.
  d. *Infection with influenza B strains before 1988*. Because sequence data were too scarce before 1988 to reliably estimate the frequencies of B/Victoria and B/Yamagata, we treated all infections before 1988 as infections with a separate “ancestral” lineage *A* and estimated protection from those infections against B/Victoria (*χ* _*VA*_) and B/Yamagata (*χ*_*YA*_). Those infections encompass the ancestral influenza B lineage before the split between B/Victoria and B/Yamagata and also strains circulating between the split and 1988. We modeled those infections to capture the age distributions of cases in people born before the 1990s, but because of the uncertain identity and antigenic phenotype of strains circulating in that period, we interpreted the associated parameters with caution.

We assumed that the protection conferred by an infection against future infections depends on the lineage but not on the time between infections. Since the lineages were identified in the late 1980s, the amino acid divergence of HA has been smaller within the lineages (≈7%) than between them (≈14%) (Figure S3; Methods: “Sequence divergence analysis”). Assuming that cross-protection from past exposures does not decay over time allowed us to calculate infection history probabilities exactly without the need for dynamical simulations (Methods: “Infection history probabilities”). Influenza vaccination coverage was low in New Zealand during the study period in the non-elderly (0.5% for children less than 1 year old, 2-5% for each of four age groups spanning 1 to 49 year-olds, and 13% for the age group 50-64 years old in 2016; Institute of Environmental Science and Research, 2017; Methods: “Demographic data”), and we thus ignored protection by vaccination.

**Table 1.** Possible infection histories in terms of the lineage of first infection and lineages encountered since.

| 1st infection | Lineages encountered later | Symbol | Susceptibility to B/Vic | Susceptibility to B/Yam |
| --- | --- | --- | --- | --- |
| None | None | $P_0$ | 1 | 1 |
| V | None | $P_{V,0}$ | $(1 - \chi_{VV})(1 - R_V)$ | $(1 - \chi_{YV})$ |
| V | Y | $P_{V,Y}$ | $(1 - \chi_{VV})(1 - R_V)$ | $(1 - \chi_{YY})$ |
| Y | None | $P_{Y,0}$ | $1 - \chi_{VY}$ | $(1 - \chi_{YY})(1 - R_Y)$ |
| Y | V | $P_{Y,V}$ | $1 - \chi_{VV}$ | $(1 - \chi_{YY})(1 - R_Y)$ |
| A | None | $P_{A,0,0}$ | $1 - \chi_{VA}$ | $1 - \chi_{YA}$ |
| A | V | $P_{A,V,0}$ | $1 - \chi_{VV}$ | $1 - \max(\chi_{YA}, \chi_{YV})$ |
| A | Y | $P_{A,Y,0}$ | $1 - \max(\chi_{VA}, \chi_{VY})$ | $1 - \chi_{YY}$ |
| A | V and Y (any order) | $P_{A,\{VY\}}$ | $1 - \chi_{VV}$ | $1 - \chi_{YY}$ |

V indicates B/Victoria, Y indicates B/Yamagata, and A indicates strains circulating before 1988. Susceptibility to each lineage depends on within-lineage ( $\chi_{VV}$ , $\chi_{YY}$ ) and cross-lineage ( $\chi_{VY}$ , $\chi_{YV}$ , $\chi_{VA}$ , $\chi_{YA}$ ) cross-protection from prior infections regardless of their order and on additional protection against the lineage first encountered ( $R_V$ and $R_Y$ ). Only the strongest cross-lineage protection term is assumed to affect susceptibility. Within-lineage protection is constrained to be stronger than cross-lineage protection against the same lineage.

We fitted the model by maximum likelihood using the probabilities of observing an infection in each cohort as the parameters of a multinomial distribution. To limit model complexity, we fitted to cases from people born since 1959, i.e., to people 60 years old or younger at the time the cases were observed. Including older people (Figure S1) would have required additional parameters to describe age-related changes in susceptibility, vaccination, healthcare-seeking behavior, and contact rates, and potentially multiple ancestral strains. We assessed the effect of moving the birth year cutoff seven years in each direction (1952 and 1966).

### Evidence for within-lineage protection and immune imprinting

The distributions of B/Victoria and B/Yamagata are most consistent with within-lineage protection against both lineages and imprinting protection against B/Yamagata, with weak or unidentifiable protection across lineages (Table 2, Figure 3, Figure S4). Any previous B/Victoria infection decreased the probability of medically attended infection with B/Victoria by 89% (*χ*_*VV*_ = 0.89, 95% CI 0.85-0.96), and any previous B/Yamagata infection decreased the probability of medically attended B/Yamagata infection by 28% (*χ*_*YY*_ = 0.28, 95% CI 0.10-0.43). The data are most consistent with weak or nonexistent protection from B/Yamagata infections against B/Victoria (*γ*_*VY*_ = 0, 95% CI 0-0.07, corresponding to a reduction in susceptibility of 0-4% after re-scaling by *χ*_*VV*_), whereas protection from B/Victoria against B/Yamagata was non-identifiable (*γ*_*YV*_ = 0, 95% CI 0-1). People for whom B/Yamagata was the lineage of first infection had an additional 87-100% reduction in susceptibility to clinical B/Yamagata infections compared with people infected with B/Yamagata after a primary infection with B/Victoria or with strains circulating before 1988 (*R*_*Y*_ = 0.96, 95% CI 0.87-1). The strength of imprinting for B/Victoria was non-identifiable (*R*_*V*_ = 0.97, 95% CI 0-1).

**Table 2.** Parameter estimates for the model fitted to the New Zealand case data.

| Parameter | MLE (95% CI) | Definition |
| --- | --- | --- |
| $\beta_1$ | 0.09 (0.07-0.10) | Baseline annual infection probability for preschoolers (0-5 years old) |
| $\beta_2$ | 0.16 (0.13-0.20) | Baseline annual infection probability for people 6-17 years old |
| $\beta_3$ | 0.12 (0.09-0.15) | Baseline annual infection probability for people 18+ years old |
| $\chi_{VV}$ | 0.89 (0.85-0.96) | Protection against B/Vic from any prior B/Vic infection |
| $\chi_{YY}$ | 0.28 (0.10-0.43) | Protection against B/Yam from any prior B/Yam infection |
| $\gamma_{YV}$ | 0.00 (0.00-1.00) | Protection from B/Vic infection against B/Yam (as a fraction of $\chi_{YY}$)<sup>†</sup> |
| $\gamma_{VY}$ | 0.00 (0.00-0.07) | Protection from B/Yam infection against B/Vic (as a fraction of $\chi_{VV}$)<sup>†</sup> |
| $R_V$ | 0.97 (0.00-1.00) | Additional B/Vic protection if 1st infection was with B/Vic |
| $R_Y$ | 0.96 (0.87-1.00) | Additional B/Yam protection if 1st infection was with B/Yam |
| $\gamma_{VA}$ | 1.00 (0.93-1.00) | Protection against B/Vic from pre-1988 infections (as a fraction of $\chi_{VV}$)<sup>†</sup> |
| $\gamma_{YA}$ | 1.00 (0.00-1.00) | Protection against B/Yam from pre-1988 infections (as a fraction of $\chi_{YY}$)<sup>†</sup> |
| $\rho$ | 1.97 (1.73-2.26) | Reporting factor for children under 2 years old. |

<sup>†</sup>Cross-lineage protection does not apply when within-lineage protection is present.

**Figure 3.**
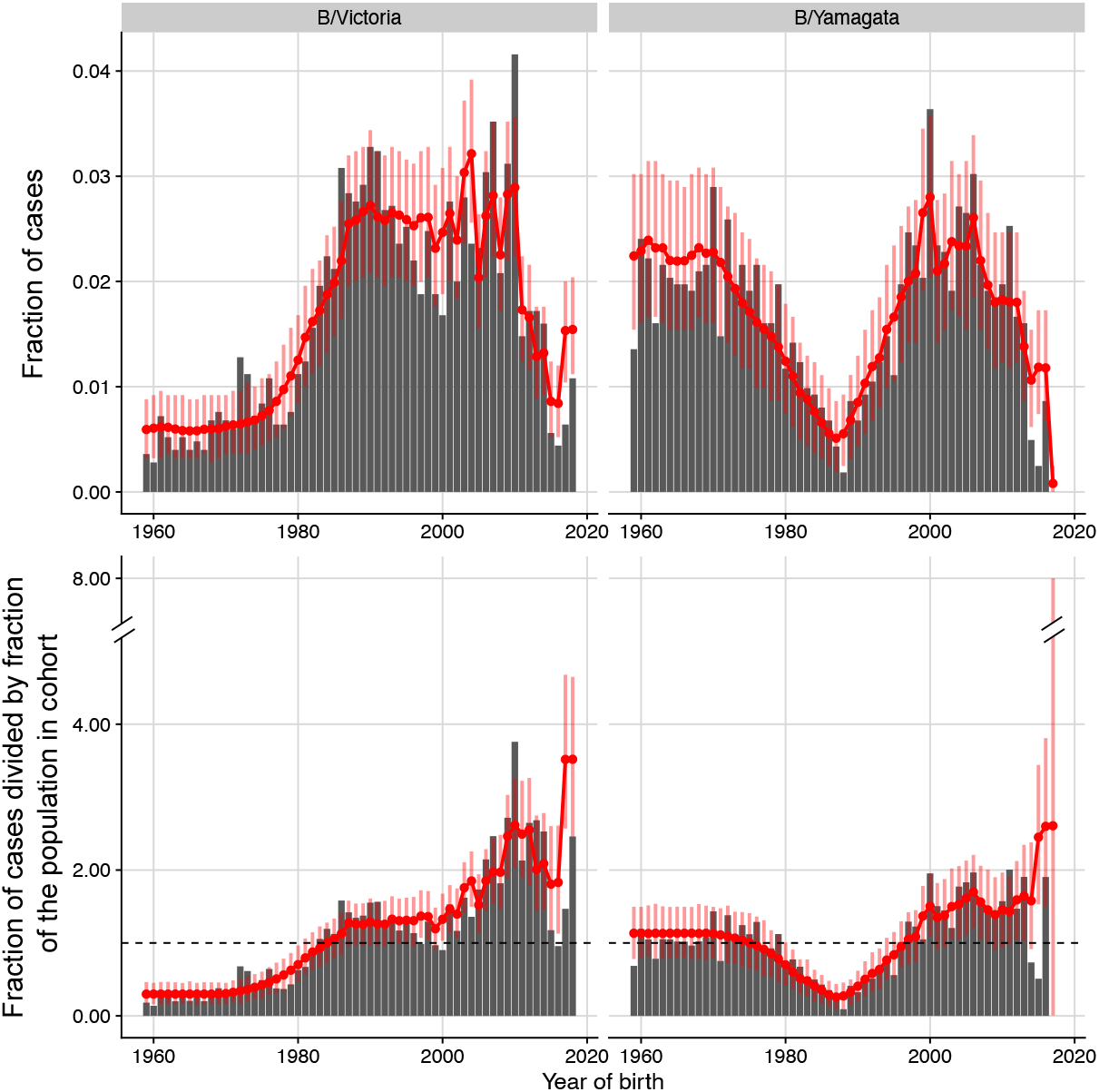
Observed and predicted distributions of influenza B cases in New Zealand by birth year. The model was simultaneously fitted to the age distributions in each observation year from 2001 to 2019, accounting for uncertainty in the birth year of each reported case given the patient’s age. For plotting, we pooled observed and predicted numbers of cases across observation years for each birth year, assuming the earliest possible birth year for each age (e.g., an age of 10 in 2000 was assumed to correspond to the birth year 1989). Red lines and dots show the predicted distribution under the model. Vertical bars are 95% bootstrap confidence intervals. In the bottom row, predicted and observed fractions of cases were normalized by dividing by the fraction of the population born in that birth year (i.e., the null expectation if all birth years were infected at the same rate).

The model suggests imprinting protection against B/Yamagata contributes to two major features of the birth year distribution of cases: the low incidence of B/Yamagata compared to B/Victoria in people born in the late 1980s and early 1990s and the lower incidence of B/Yamagata in those birth cohorts compared with older cohorts (Figure 3). Because B/Yamagata appears to have been the only lineage circulating in New Zealand in the 1990s, many more people born in the late 1980s and early 1990s had been infected with B/Yamagata than with B/Victoria by the time the cases were recorded in the 2000s and 2010s (Figure 4). Under the maximum likelihood estimates of infection history probabilities, 87% of people born between 1987 and 1993 had been infected with B/Yamagata by 2010 (the midpoint of the surveillance period), of which 96% had B/Yamagata as their first influenza B infection (Figure 4). In contrast, only 55% of people in those birth cohorts had been infected with B/Victoria by 2010. Thus, cohorts born in the late 1980s and early 1990s were more susceptible to B/Victoria (about 50% as susceptible as a fully naive cohort) than to B/Yamagata (20% as susceptible).

**Figure 4.**
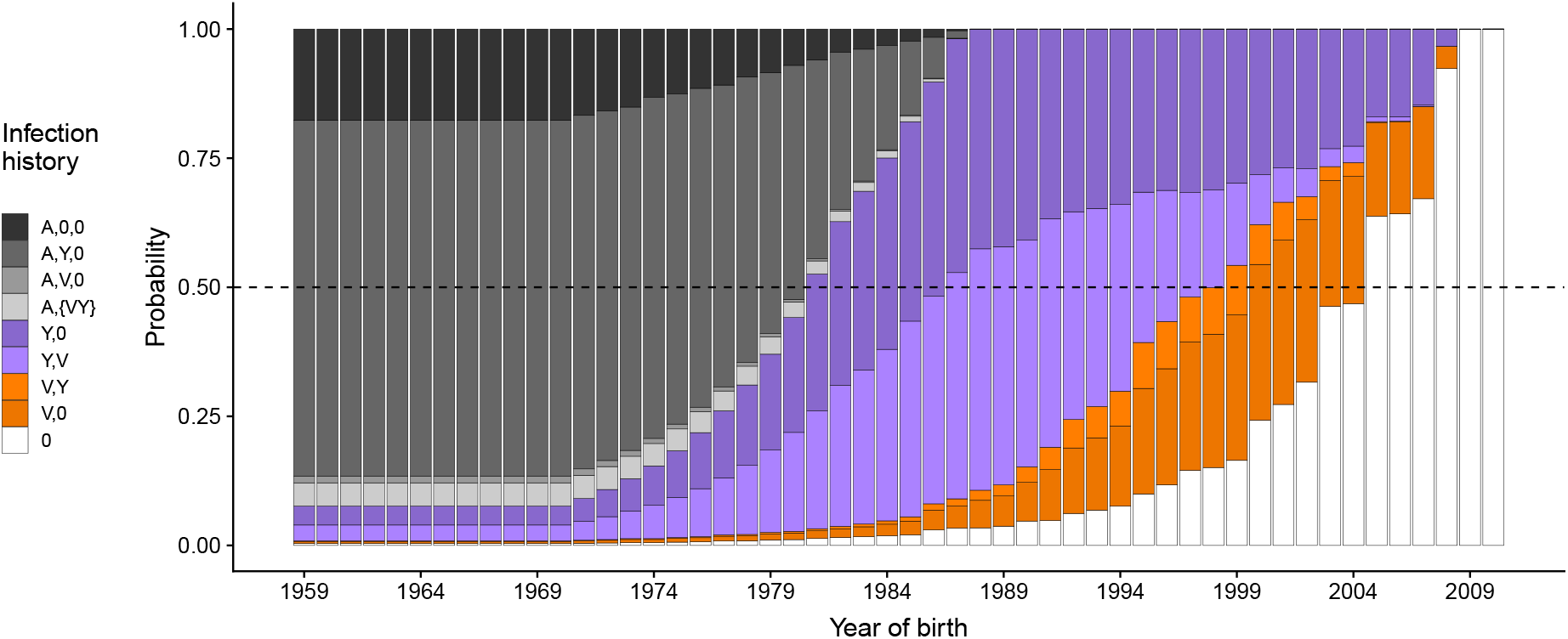
Probabilities of different infection histories with influenza B in New Zealand for people born between 1959 and 2010 and observed in 2010. Infection histories consist of the lineage of first infection and lineages encountered later regardless of their order. Probabilities were estimated by fitting the model to case data from New Zealand. (A,0,0): First infection before 1988 and no subsequent infections with either B/Victoria or B/Yamagata. (A,Y,0) and (A,V,0): First infection before 1988 followed by B/Yamagata but not B/Victoria and by B/Victoria but not B/Yamagata, respectively. (A,{V,Y}): First infection before 1988 followed by infections with both B/Victoria and B/Yamagata in any order. (Y,V) and (Y,0): First infection with B/Yamagata, with and without a subsequent B/Victoria infection. (V,Y) and (V,0): First infection with B/Victoria, with and without a subsequent B/Yamagata infection. (0): fully naive to influenza B.

Older birth cohorts, in contrast, were also likely infected with B/Yamagata in the 1990s and 2000s (74% of people born until 1986 had been infected with B/Yamagata by 2010), but unlike younger cohorts, they lacked the imprinting protection against B/Yamagata (Figure 4). The high proportion of B/Yamagata cases in those older birth cohorts in the 2000s and 2010s, even after many people in them had already been infected with B/Yamagata, is consistent with only moderate within-lineage protection against B/Yamagata in the absence of immune imprinting. The difference in the proportion of B/Yamagata cases between cohorts born around 1990 and older birth cohorts also appears not due to differences in transmission related with age itself, since we estimated similar baseline annual infection rates for school-age children and adults (Table 2).

Weak or nonexistent protection from B/Yamagata against B/Victoria is consistent with a surge of B/Victoria cases in children born in the late 1980s and early 1990s as B/Victoria reemerged in New Zealand in the early 2000s (Figure S5). By 2000, 66% of people born in 1987-1993 had been infected with B/Yamagata. The high incidence of B/Victoria in those birth cohorts in the early 2000s suggests their history of B/Yamagata exposure did not protect them against medically attended B/Victoria infections. This lack of protection from B/Yamagata against B/Victoria was distinguishable from potential changes in transmission with age (Figure S6). The non-identifiability of cross-lineage protection in the other direction, from B/Victoria against B/Yamagata, might be due to the model estimating that relatively few people were infected with B/Victoria alone (Figure 4). Our model assumes that protection from B/Yamagata infections themselves supersedes the protection from B/Victoria infections against B/Yamagata in people infected with both lineages.

The estimated protection from strains circulating before 1988 against clinical infections suggests those early strains may have been antigenically more similar to B/Victoria than to B/Yamagata. Protection against B/Yamagata from strains circulating before 1988 (relative to protection from B/Yamagata itself) was non-identifiable (*γ*_*YA*_ = 1.00, 95% CI 0.00-1.00, corresponding to a 0-28% protection). It is possible that protection from those early strains against B/Yamagata violates our assumption that it cannot be stronger than protection from B/Yamagata itself, but the high proportion of B/Yamagata cases throughout the 2000s and 2010s in birth cohorts that were likely infected with those early strains suggests that such protection is limited at best (Figure S5). In contrast, the model estimated that infections with early strains were as protective against B/Victoria as were B/Victoria infections themselves (*γ*_*VA*_ = 1.00, 95% CI 0.93-1.00). Thus, when B/Victoria began circulating at high frequencies in the 2000s, birth cohorts with a history of influenza B infection prior to 1988 were strongly protected against medically attended B/Victoria infections.

### Sensitivity analyses corroborate the exclusive circulation of B/Yamagata in New Zealand in the 1990s

While previous work suggests B/Yamagata was the only lineage circulating widely outside of East Asia in the 1990s (Shaw et al., 2002), the evidence for the exclusive circulation of B/Yamagata in New Zealand during that period is limited. Few isolates collected in New Zealand or Australia during the 1990s were deposited in sequence databases (although they are all B/Yamagata, Figure 1b, Figure S2), and antigenic surveillance often did not test circulating strains against representative B/Victoria strains. We re-fitted the model using lineage frequencies estimated from all countries represented in sequence databases for the 1990s — resulting in an average B/Victoria frequency of about 25% in that period (Figure S7) — and found that the age distributions of cases could not be adequately fitted using those alternative frequencies (a decrease of 53 log-likelihood units compared to the model fitted with only B/Yamagata in the 1990s for the same number of parameters). In particular, even with strong imprinting and cross-lineage protection from B/Victoria, the model could not fully capture the low number of B/Yamagata cases in cohorts born around 1990 (Figure S8). Thus, the age distributions of medically attended influenza B cases in New Zealand are most consistent with the exclusive circulation of B/Yamagata in the 1990s.

Because we grouped infections before the lineages split with infections that occurred between the split and 1988, it is possible that the strong protection from those early strains against B/Victoria derives from a high incidence of B/Victoria during that period (which we are unable to detect). This potential period of B/Victoria dominance might also lead to imprinting protection against B/Victoria in cohorts born then. To investigate this possibility, we fitted the model assuming B/Victoria was the only lineage circulating from 1983 to 1990 and found a poorer fit to the data (by about 6 log-likelihood units) but overall similar estimates of protection (Figure S9). Even assuming this period was dominated by B/Victoria, the data did not conclusively support imprinting protection against it (*R*_V_ = 0, 95% CI 0-0.43). Cohorts born before the lineages split in the early 1980s do not appear more susceptible to B/Victoria than those born between the lineage split and 1990, when B/Victoria might have dominated (Figure 3). We also simulated data identical in structure to the New Zealand case data to verify that if B/Victoria had strong imprinting protection but limited within-lineage protection, as estimated from the observed data for B/Yamagata, the model would have correctly identified that scenario (Figure S10).

### The model captures broad features of the age distributions of cases in other regions

If the estimated strengths of within- and cross-lineage protection are correct, our model should be able to explain the age distributions of cases outside New Zealand. Since additional data sets with clear sampling criteria were not available, we used the model to predict the age distributions of influenza B sequences from the European Union (EU) and China on the GISAID database and the age distribution of isolates from Australian hospital samples compiled by Vijaykrishna et al. (2015) (Methods: “Age distributions of cases in non-surveillance data”). In the absence of clear sampling criteria, the age distributions in those data sets might be affected by under or oversampling of particular age groups or by changes in sampling over time, factors that were not included in our model.

Despite those potential biases, and despite uncertainty in historical lineage frequencies in Europe and East Asia, the model reproduced major features of the age distributions in the additional data sets using estimates of protection from New Zealand (Figure S11). The model correctly predicted fewer B/Yamagata cases than B/Victoria cases in cohorts born around 1990 in the EU and Australia, where, as in New Zealand, only B/Yamagata circulated in the mid to late 1990s (all 51 isolates from 1995 to 1999 were B/Yamagata; Figure S2). In contrast, B/Victoria appears to have never stopped circulating during the 1990s in East Asia (Shaw et al., 2002; Figure S2), although B/Yamagata might have been more common (65% of 275 isolates from East Asia in the 1990s). Compared to the other regions, the continued circulation of B/Victoria in East Asia would have decreased the difference in immunity against B/Yamagata versus B/Victoria in cohorts born in the 1990s. Accordingly, the proportions of B/Victoria and B/Yamagata cases in those cohorts were more similar in China than in the other regions. This smaller difference was partly predicted by the model (Figure S11). However, the model still predicted a high proportion of B/Yamagata cases in older cohorts, which was not apparent in the Chinese data. When we re-estimated parameters separately for each data set, imprinting protection against B/Yamagata was inferred to be strong in the European and Australian data but was not identifiable in the Chinese data (Table S1, Figure S12). Support for imprinting protection against B/Yamagata in the European data was robust to uncertainty in lineage frequencies in the early 1990s, when scarce isolate data suggest B/Victoria, and not B/Yamagata, may have dominated in Europe (Figure S2, Table S1).

Estimates of cross-lineage protection in these additional datasets differed from the New Zealand estimates. This discrepancy might reflect differences in severity or health-seeking behavior between the data sets, for instance if cross-lineage protection occurs for severe cases but not milder cases. In contrast to the other data sets, the model estimated perfect within-lineage protection against B/Yamagata in China to account for the small proportions of B/Yamagata cases in older cohorts. This result too might arise from differences in severity between data sets, for instance if older cohorts are susceptible to mild B/Yamagata cases but protected against severe ones. However, our model cannot disentangle those effects from sampling biases.

### Estimated infection probabilities are consistent with independent serological data

To test if our model produced realistic estimates of infection probabilities given complete susceptibility (*β*_*i*_ parameters), we compared estimates from the model fitted to the New Zealand case data with estimates based on cross-sectional serology from children in the Netherlands (Bodewes et al., 2011; Methods: “Model validation with independent serological data”). Both the infection probabilities estimated from our model and those inferred from cross-sectional serology represent an average probability across influenza seasons. Despite potential differences between New Zealand and the Netherlands in the intensity of influenza B circulation and in lineage frequencies, we found similar estimates of infection risk. The annual probability of influenza B exposure was similar between our model (9% for preschoolers and 16% for school-age children, 95% CIs 7-10% and 13-20%) and the estimates from Dutch seroprevalence data (12% for preschoolers and 22% for school-age children, 95% CIs 10-14% and 16-27%). These estimates are also within the range of infection probabilities estimated from longitudinal studies (Glezen et al., 1980; Frank et al., 1983; Monto et al., 1985; Horby et al., 2012; Hayward et al., 2014; Huang et al., 2019). The fraction of children with detectable antibodies against B/Victoria was close to the prediction from our model, but more children in the Netherlands had detectable B/Yamagata antibodies than predicted by the model for New Zealand (Figure S13). In addition to differences in lineage frequencies between New Zealand and the Netherlands, this discrepancy might be due to the presence of antibodies from B/Victoria infections that cross-react with B/Yamagata but not vice-versa (Rota et al., 1990; Levandowski et al., 1991a,b; Skowronski et al., 2012; Laurie et al., 2018).

We performed two sensitivity analyses to test if our conclusions held if the model were not given the freedom to estimate baseline infection probabilities (*β*_*i*_) and differential reporting for children under 2 years old (*ρ*). First, we fitted the model while removing the age-specific reporting parameter and constraining the baseline exposure probabilities to the values inferred from the Dutch seroprevalence data (using 22% for both school-age children and adults, since adults were not represented in those data). Second, we fitted the model while constraining baseline infection probabilities to be 30%, as reported by the Houston Family Study (Glezen et al., 1980; Frank et al., 1983), representing the upper range of independent estimates. Despite poorer fits to the data, parameter estimates were similar to those of the main analysis in both cases, except for protection from B/Victoria against B/Yamagata (*γ*_*YV*_ 95% CI 0.02-0.66 and 0-0.38, compared with 0-1 in the main analysis; Figure S14, Figure S15). Under a 30% exposure probability, the data would also support at most a very small imprinting protection against B/Victoria (*R*_*V*_ 95% CI 0-0.07, compared to 0-1 in the main analysis). Thus, our main conclusions were independent of the model’s ability to estimate baseline exposure probabilities and differential reporting in children.

### Additional sensitivity analyses

Additional sensitivity analyses showed that estimates of protection based on the New Zealand data were similar if we excluded non-surveillance cases (Figure S16, Figure S17), if we moved the minimum birth year to which we fitted the model (1959) by 7 years in each direction (Figure S18, Figure S19), and if we assumed a separate reporting parameter for children under 5 years old instead of children under 2 years old (Figure S20). Fitted to both surveillance and non-surveillance cases combined, the model underestimated cases in the most recent birth cohorts due to variation over time in the number of non-surveillance cases reported in the youngest children (Figure S5). This variation might have been caused by changing sampling practices, for instance if hospital samples from children were more likely to be collected in some years. Fitted to surveillance cases alone, however, the model predicted the number of cases in these birth cohorts well (Figure S17). Estimates of the remaining parameters were similar if we fixed at zero the two parameters that had large point estimates but were completely non-identifiable (imprinting protection against B/Victoria and protection from strains circulating before 1988 against B/Yamagata; Figure S21).

Finally, because in children the first influenza infection also tends to be a recent infection, estimates of immune imprinting based on case data that include children might reflect strong protection against recent infection instead of the effect of early infections *per se* (Arevalo et al., 2020b). To investigate this possibility, we re-fitted the model after excluding cases in children under 10 years old and found similar results (Figure S22). Thus, our estimate of B/Yamagata imprinting likely reflects the effect of early exposures.

## Discussion

Our model suggests that differences in the age distributions of B/Victoria and B/Yamagata cases can be explained by historical changes in lineage frequencies combined with cross-protection between strains of the same lineage and additional protection against B/Yamagata in people first infected with it. In particular, two major features of the age distributions seem to arise from the dominance of B/Yamagata in New Zealand in the 1990s. First, the model suggests that people born in the late 1980s and early 1990s had fewer B/Yamagata cases than B/Victoria cases in the 2000s and 2010s because they had accumulated immunity against B/Yamagata but not B/Victoria during the 1990s. Second, imprinting of people born in the late 1980s and early 1990s with B/Yamagata led to fewer B/Yamagata cases in those birth cohorts than in older and younger ones, whose primary influenza B infections were less likely to be B/Yamagata.

While previous work revealed immune imprinting at the level of the HA groups and subtypes of influenza A (Gostic et al., 2016, 2019; Arevalo et al., 2020b), our results suggest a similar phenomenon occurs between more recently diverged and thus antigenically closer branches of influenza B. The major antigens HA and NA evolve more slowly within the influenza B lineages than within influenza A subtypes (Bedford et al., 2014, 2015; Vijaykrishna et al., 2015), and the amino acid sequence divergences are much lower between B/Victoria and B/Yamagata (≈14%, 2% and 7% in 2019 for the HA head, the HA stalk, and NA, respectively) than between H1N1 and H3N2 (≈66%, 50% and 60%, respectively) (Figure S23). Still, epitopes conserved within the lineages but variable between them (or between the lineages and their ancestor) might be the basis for imprinting protection against B/Yamagata (and potentially B/Victoria), as has been hypothesized for influenza A subtypes (Gostic et al., 2019; Arevalo et al., 2020a,b; Carreño et al., 2020).

While imprinting protection against B/Yamagata has a clear effect on the age distributions of cases, the non-identifiability of imprinting protection against B/Victoria might be due to the lack of an extended period when B/Victoria circulated at consistently higher frequencies than B/Yamagata. Our analysis is limited by the scarcity of data on the identity and antigenic phenotype of influenza B strains circulating prior to the lineage split and on lineage frequencies shortly after. However, even if we assumed B/Victoria circulated alone between the lineage split and 1990, the data did not conclusively support imprinting protection against B/Victoria. We speculate that antibodies elicited by primary B/Yamagata infections might target epitopes that are more conserved (or affect viral neutralization more strongly) than epitopes targeted by antibodies from primary B/Victoria infections.

The model suggests protection from B/Yamagata against medically attended B/Victoria infections is weak at most, but it could not estimate the strength of cross-lineage protection in the other direction. Previous serological and vaccine effectiveness studies disagree on the strength of cross-lineage protection. B/Yamagata often induces low or undetectable levels of HA antibodies cross-reactive to B/Victoria (Rota et al., 1990; Levandowski et al., 1991a,b; Skowronski et al., 2012; Laurie et al., 2018), consistent with our estimate of weak protection. The same studies of strains from the late 1980s, 2000s and 2010s show that exposure to B/Victoria induces antibodies that inhibit hemagglutination by B/Yamagata, suggesting B/Victoria might protect against B/Yamagata even if this protection does not strongly affect the age distributions of cases. In contrast to the asymmetric cross-lineage protection suggested by these serological observations, cross-lineage protection in both directions has been observed in some vaccine efficacy studies but not in others, with unexplained variation across seasons and age groups (Belshe et al., 2010; Tricco et al., 2013; Ohmit et al., 2014; McLean et al., 2015; Skowronski et al., 2019; Drori et al., 2020; Gaglani et al., 2020). Longitudinal studies of infection and vaccination might reveal if cross-lineage protection differs between infection and vaccination or varies with age (e.g., via increased bias toward conserved epitopes in adults; Nachbagauer et al., 2016; Sun et al., 2019) or vaccine type (inactivated or live attenuated). Studies of children first exposed to influenza B via vaccination might also reveal if imprinting can occur via trivalent vaccines containing only one influenza B lineage or quadrivalent vaccines containing both.

Our results also suggest that the acquisition of a B/Yamagata NA gene by B/Victoria strains via reassortment in 2000-2001 (Dudas et al., 2015; Langat et al., 2017) did not lead to substantial protection against the reassortant strains in people previously infected with B/Yamagata. While most studies of the antibody response to influenza focus on HA, serological evidence suggests immune responses targeting NA but not HA are common for influenza B, especially in children (Huang et al., 2019). Yet birth cohorts with a history of B/Yamagata infections had a high proportion of clinical B/Victoria infections in the 2000s, even though all B/Victoria cases in the data occurred after B/Victoria had acquired a B/Yamagata NA. These results suggest that protection against B/Victoria is not strongly mediated by anti-NA antibodies.

The difference in the estimated strength of within-lineage protection without imprinting for B/Victoria (85-96%) and B/Yamagata (10-43%) might be due to differences in the times of infection for the birth cohorts informing each of the estimates and different rates of antigenic evolution between the lineages. Because most people born since the late 1980s and infected with B/Yamagata were imprinted with it, our estimate of within-lineage protection against B/Yamagata in the absence of immune imprinting is mostly informed by cases in older birth cohorts lacking imprinting protection against B/Yamagata. Most of the B/Yamagata infections in those birth cohorts would have occurred in the 1990s, when B/Yamagata was dominant. In contrast, our estimate of within-lineage non-imprinting protection against B/Victoria was informed by more recent B/Victoria infections in younger birth cohorts, including people who were imprinted with B/Yamagata in the 1990s and later infected with B/Victoria in the 2000s. Because these B/Victoria infections were more recent, they were potentially antigenically closer to modern strains than were the B/Yamagata infections that inform estimated within-lineage protection against B/Yamagata, and thus less likely to violate our assumption of constant within-lineage protection over time. However, B/Victoria appears to undergo slightly faster antigenic evolution than B/Yamagata (Bedford et al., 2014; Vijaykrishna et al., 2015), with several clades containing amino acid deletions emerging since 2015 (Virk et al., 2020). These deletions have not been accompanied by an increase in B/Victoria cases in cohorts in which past B/Victoria infections are common (Figure S5), suggesting they did not strongly affect within-lineage protection against B/Victoria. Longitudinal studies of infection risk might reveal if within-lineage protection against B/Yamagata without imprinting is stronger in recent birth cohorts.

It remains unclear precisely how differences in people’s antibody responses and past infections shape their susceptibility to influenza. Our model and others (Gostic et al., 2016, 2019; Arevalo et al., 2020b) show how analyses based on birth cohorts can reveal broad dynamics and potential mechanisms of immune protection. Characterizing antibody specificity after infection and vaccination might reveal the mechanisms behind these patterns. Antibody targeting of NA and of particular HA epitopes is known to change with age (Nachbagauer et al., 2016; Rajendran et al., 2017; Sun et al., 2019), but few studies have linked antibody specificity to the infection history of particular birth cohorts and their susceptibility to particular variants (Linderman et al., 2014; Andrews et al., 2015; Petrie et al., 2016). This resolution could improve our understanding of influenza epidemiology and the response to influenza vaccination.

## Methods

### Case data

Medically attended influenza B cases in New Zealand were identified from samples taken from patients with influenza-like illness (ILI) attended by a network of general practitioners recruited for surveillance (2,430 cases with an identified influenza B lineage) and from non-surveillance hospital samples (1,606 cases with an identified lineage) analyzed by regional diagnostic laboratories and by the WHO National Influenza Centre at the Institute for Environmental Science and Research (ESR). Briefly, general practice surveillance operates from May to September, with participating practices collecting nasopharyngeal or throat swabs from the first ILI patient examined on each Monday, Tuesday and Wednesday. ILI is defined as an “acute respiratory tract infection characterised by an abrupt onset of at least two of the following: fever, chills, headache and myalgia” (Huang et al., 2008). A subset of the New Zealand data (cases from 2002-2013) was previously compiled by Vijaykrishna et al. (2015) along with cases from Australia reported to the World Health Organization (WHO) Collaborating Centre for Reference and Research on Influenza in Melbourne.

### Statistical model of influenza B susceptibility based on infection history

For lineage *V* (B/Victoria), we modeled the number of cases in people born in birth year *b* observed in season *y* as a multinomial draw with probabilities given by:

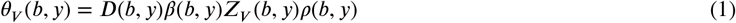

with an analogous equation defining the multinomial distribution *θ*_*Y*_ (*b, y*) for lineage *Y*(B/Yamagata). *D*(*b, y*) is the fraction of the population that was born in year *b* as of observation season *y. Z*(*b, y*) is the susceptibility to lineage *V* during season *y* of a person born in year *b* relative to that of an unexposed person. *β* (*b, y*) is a baseline probability of infection with influenza B that captures differences in transmission associated with age (thus depending on *b* and *y*) and is equal to *β*_1_ if people born in year *b* are in preschool during season *y* (0-5 years old), *β*_2_ if they are school-age children or teenagers (6-17 years old), or *β*_3_ if they are 18 or older. *ρ* (*b, y*) is an age-specific factor equal to a parameter *ρ* if people are less than 2 years old and 1 otherwise. While *β* (*b, y*) represents true differences in infection probabilities between preschoolers and post-preschool individuals and thus affects the infection history probabilities in the calculation of *Z*(*b, y*), *ρ* (*b, y*) represents a differential probability of receiving medical attention and does not affect those probabilities. For each lineage and observation season, values of *θ* from Eq. (1) are normalized by their sum across birth years to make them proper multinomial probabilities.

We defined relative susceptibility to *V, Z*_*V*_ (*b, y*), as an expectation over all possible immune histories in terms of the lineage of first infection and subsequent infection with the other lineage. We let susceptibility be 1 for a person never exposed to influenza B. Cross-immunity from any previously encountered strain of *V* or *Y* decreased susceptibility to the corresponding lineage by *χ*_*VV*_ and *χ*_*YY*_, respectively. Susceptibility was further reduced by *R*_*V*_ or *R*_*Y*_ if the lineage of first infection was *V* or *Y*. In the absence of a previous homologous infection, previous infection with *Y* decreased susceptibility to *V* by *χ*_*VY*_, and previous infection to *V* decreased susceptibility to *Y* by *χ*_*YV*_. Protection due to homologous infections superseded cross-lineage protection. Protection against *V* from a previous *Y* infection was constrained to be a fraction of the within-lineage protection to *V* (and vice-versa): *χ*_*VY*_ = *χ*_*VV*_ · *γ*_*VY*_, *χ* _*YV*_ = *χ*_*YY*_ · *γ*_*YV*_, where 0 ≤ *γ*_*VY*_, *γ*_*YV*_ *≤* 1. Similarly, pre-1988 infections reduced susceptibility to *V* by *χ*_*VA*_ = *χ* _*VV*_ · *γ*_*VA*_ and to *Y* by *χ*_*YA*_ = *χ*_*YY*_ · *γ*_*YA*_. Finally, *Z*_*V*_(*b, y*) was calculated as the sum of susceptibilities in Table 1 weighted by the probabilities of the corresponding infection histories (below). Relative susceptibility to *Y, Z*_*Y*_(*b, y*) was defined analogously.

We estimated parameters by maximum likelihood using R (version 3.4.3) and package optimParallel. We calculated the total likelihood as the product of the likelihood for each lineage in each observation year, with the number of cases in each combination treated as an independent multinomial draw. For plotting, we summed the observed and predicted cases for each birth year across observation years.

### Infection history probabilities

To calculate the probabilities in Table 1, we assumed infections occur in discrete time measured in units of annual influenza seasons. We considered possible infections in each season between birth and the last season before observation season *y*. For a person born in year *b* and previously unexposed to influenza B, we let *a*_*b,i*_ be the probability of becoming infected in season *i*. This probability was then modified by protection from previous infections as in Table 1. Given that an infection occurred in season *i*, we assumed that the probability it was caused by lineages *A* (“ancestor”), *V* or *Y* was equal to their frequencies in that season, *f*_*A,i*_, *f*_*V,i*_ and *f*_*Y,i*_, with *f*_*A,i*_ = 1 for all *i* before 1988, and *f*_*V,i*_ + *f*_*Y,i*_ = 1 since (Figure 1). For simplicity, we assumed that people could not be infected more than once in each season (including simultaneous infections by the two lineages.)

Let 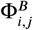 be the probability that no infections with influenza B occurred for a naive person born in *b* from seasons *i* to *j* (inclusive). It is given by

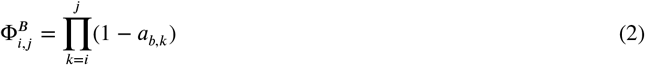

where *k* indexes years (influenza seasons). Thus the first probability in Table 1, that of being fully naive to influenza B, is given by:

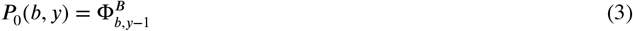

To shorten the expressions for the remaining probabilities, we let 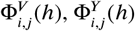 and 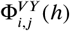 be the probability that no *V* infections, no *Y* infections, and neither *V* nor *Y* infections occurred between seasons *i* and *j* (inclusive), respectively. Unlike Φ^*B*^, which applies to naive people, Φ^*V*^, Φ^*Y*^ and Φ^*VY*^ depend on the person’s infection history, *h*. To calculate the probability of an initial infection with *A* but no subsequent infections with *V* or *Y, P*_*A*,0,0_(*b, y*), we integrated, across all possible seasons *i* of first infection, the joint probability that the person’s first influenza B infection occurred in season *i* with the ancestral lineage *A* and that no subsequent infections with *V* or *Y* occurred from *i* + 1 to *y* − 1:

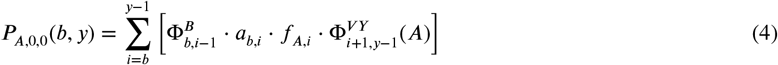

where the probability that the first infection with influenza B occurred in season *i* with the ancestor *A* is obtained by multiplying the probability of no previous infections 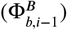 by the probability of an infection in season *i* (*a*_*b,i*_) and by the frequency of lineage *A* in season *i* (*f* _*A,i*_). The probability of no infections with either *V* or *Y* after the initial infection with *A* in season *i*, 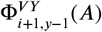, depends on protection from the *A* infection against *V* (*χ*_*VA*_) and *Y* (*χ*_*YA*_):

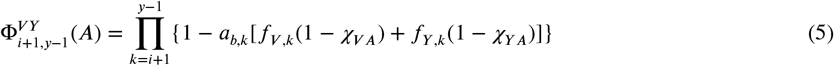

Similarly, the joint probability of first infection with the ancestor *A* and subsequent infection with *V*, but not with *Y*, is given by

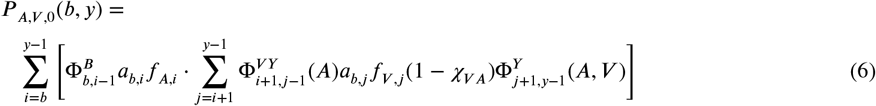

where we again integrated over all possible seasons *i* when the first infection with influenza B might have occurred. Given the first infection occurred in season *i* with the “ancestral” lineage *A*, we calculated the probability of subsequent infection with *V*, but not with *Y*, by integrating over all possible seasons *j* when the first infection with *V* may have occurred. Given the initial infection with *A* in season *i*, the joint probability that the first *V* infection occurred in *j* is given by the probability that neither *V* nor *Y* infections occurred from *i* + 1 to 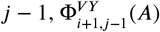, times the probability of a *V* infection in season *j*, given by *a*_*b,j*_ *f*_*V,j*_(1 − *χ*_*VA*_). The probability 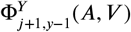 of no subsequent *Y* infections after season *j* given the previous *A* and *V* infections is then given by

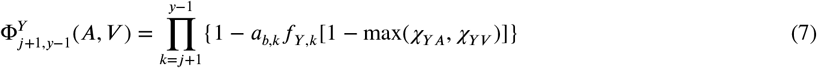

where we assumed that only the strongest cross-protection (from the previous *A* and *V* infections) applies. By analogy, for *P*_*A, Y*,0_(*b, y*) we have

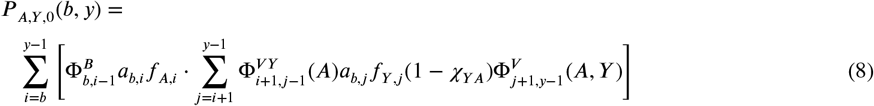

where

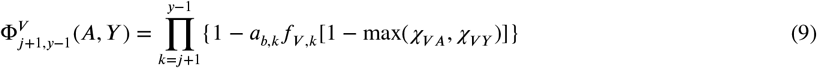

To calculate *P*_*A*,{*VY*}_(*b, y*), we first computed the probabilities for the particular cases where either *V* or *Y* were the second infection, *P*_*A, V→Y*_ (*b, y*) and *P*_*A, Y→V*_ (*b, y*), such that *P*_*A*,{*VY*}_(*b, y*) is the sum of the two. The first is given by

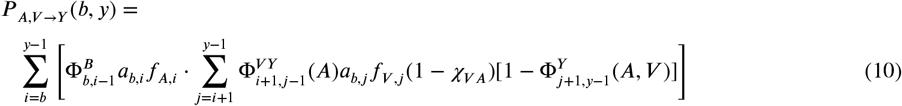

and the second is given by

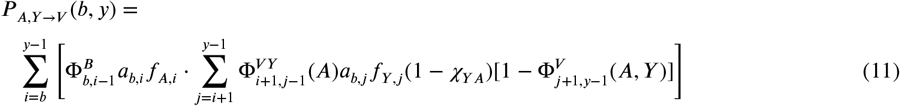

Next we write down the probabilities of infection histories with either *V* or *Y* as first infections and no subsequent infections. For *P*_*V*, 0_(*b, y*):

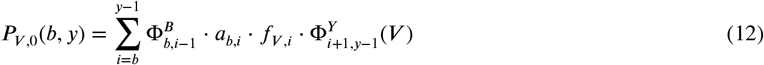

where

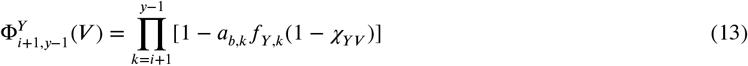

By analogy, for *P*_*Y*, 0_(*b, y*)

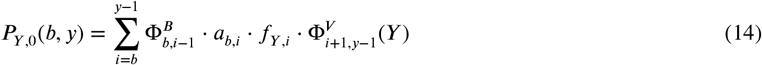

where

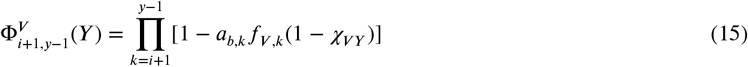

Finally, *P*_*V,Y*_ (*b, y*) and *P*_*Y,V*_ *(b, y)* are given by:

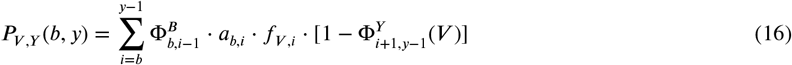

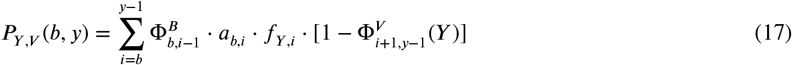

Because case data had information on age and not the exact birth year, we averaged each exposure history probability across the two possible birth years given the age and the observation year (for instance, a 10-year-old in 2000 may have been born in either 1989 or 1990). Because the probabilities of different infection histories become very similar for cohorts born long before the lineages split, we used the probabilities calculated for the birth year 1970 for all previous cohorts to decrease computation time.

### Season-specific attack rates

Let P_inf_(*b, i, t*) be the probability that a previously unexposed person born in year *b* has been infected with influenza B after experiencing fraction *t* ∈ [0, 1] of season *i*. Assuming a constant instantaneous attack rate *α*_*b,i*_ throughout the season, P_inf_(*b, i, t*) is given by:

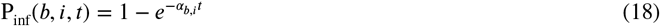

We let the instantaneous attack rate *α*_*b,i*_ be equal to an age-specific baseline multiplied by an intensity score *S*_*i*_ representing the strength of influenza B circulation in season *i* relative to other seasons:

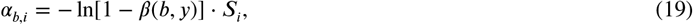

where *β* (*b, y*) takes on value *β*_1_, if birth year *b* corresponds to an age of less than 5 in year *y, β*_2_, if the corresponding age is 6-17, and *β*_3_, for ages 18 and older. The probability of infection for an unexposed person born in year *b* across the entire season, *a*_*b,i*_, is obtained by substituting *α*_*b,i*_ in Eq. [18] and setting *t* = 1:

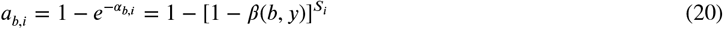

The definition of *α*_*b*_,_*i*_ in Eq. (19) was chosen such that for a season with average influenza intensity (*S*_*i*_ = 1), the annual probability of infection for an unexposed person is equal to *β* (*b, y*).

For the season corresponding to the first year of life (*i* = *b*), people are only susceptible to infections during a fraction of the season, depending on when they were born and how long they were protected by maternal antibodies. In those cases, we defined *a*_*b,i*_ as the expected probability of infection across all possible weeks of birth:

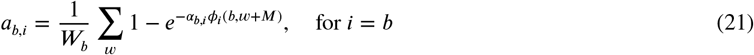

where *ϕ*_*i*_(*b, w*) is the fraction of cases in season *i* observed in or after week *w* of year *b* and *W*_*b*_ is the number of weeks in year *b*. Because people are assumed to be completely protected against infection by maternal antibodies for the first *M* weeks following birth, *ϕ*_*i*_ was computed for an effective birth week *w* + *M*. Based on the fraction of children under the age of 1 with detectable antibodies to influenza B (Bodewes et al., 2011), we set *M* to 26 weeks (approximately 6 months). Averaging *ϕ*_*i*_(*b, w* + *M*) over all possible birth weeks *w* in year *b* gives the expected fraction of season *i* experienced by a person born in year *b* assuming births are distributed uniformly in time. We estimated *ϕ*_*i*_(*b, w*) by fitting the incomplete beta function to the cumulative fraction of cases in the seasons for which we had case data, using R package FlexParamCurve. Following Gostic et al. (2016), we truncated season-specific infection probabilities so that they never exceed 0.75 even in years of high estimated influenza B intensity.

### Intensity scores

We defined the intensity score *S*_*i*_ in Eq. (19) as:

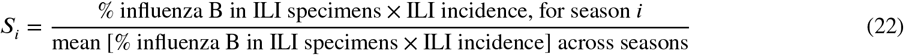

Annual influenza surveillance reports from New Zealand’s Institute of Environmental Science and Research (ESR) available from 2003 on (Institute of Environmental Science and Research, 2020) give the “isolation” or “detection” rate (the number of influenza-positive swabs divided by the number of swabs tested), the percentage of influenza A and B viruses among all influenza-positive isolates (both “sentinel” and “non-sentinel”), and the estimated number of ILI cases in New Zealand for each season. The reports do not directly give the fraction of ILI specimens that were influenza B-positive. Instead, we calculated the fraction of ILI isolates that were influenza B-positive in a season by multiplying the fraction of ILI isolates that were influenza-positive by the fraction of influenza-positive specimens that were influenza B. For seasons without data on the fraction of influenza-positive specimens in ILI specimens (1988 to 2000), without data on the fraction of influenza B in influenza-positive specimens (1988-1989), or without estimates of the total number of ILI cases (1988-2001), we used the average values of those quantities across the remaining seasons. Although reports are not available for 1990-2002, the 2003 annual report lists the frequency of influenza B in influenza-positive specimens for those seasons.

The World Health Organization’s FluNet has weekly data on the fraction of ILI specimens that were influenza B-positive in Australia from 1997 to the present. However, in data from before 2003, the number of influenza-positive specimens was usually the same as the reported number of specimens processed for that week, suggesting strong case ascertainment or reporting bias. We thus used data on the percent of influenza-positive ILI cases from 2003 on. Annual influenza reports from 1994 to 2010 are available from the Australian government’s Department of Health website (Australian Government Department of Health, 2019). They report numbers of influenza A- and B-positive isolates but not the total number of specimens tested. Thus, only the fraction of influenza B in influenza isolates (but not in ILI isolates) can be estimated from those reports. We therefore used data from the WHO to calculate the fraction of influenza B-positive specimens in ILI specimens for 2003-2017. For 1994-2002, we multiplied the fraction of influenza B in influenza-positive specimens for each season (from the Department of Health reports) by the average annual fraction of influenza-positive specimens in ILI specimens from 2003-2017 (from the WHO data) to arrive at the fraction of influenza B positive ILI specimens. Finally, for seasons where data were missing altogether (1988-1993), we used the average annual fraction of influenza B positive specimens in ILI specimens for subsequent seasons (1994-2017).

To estimate ILI incidence in Australia, we used the maximum weekly number of ILI cases per 1000 consultations for each season from Department of Health annual and weekly reports (weekly reports are available for years since the last annual report in 2010). Different ILI definitions were used from 1994-2003 and from 2004-2010, and starting in 2009 reported weekly ILI rates were averaged from multiple branches of the Australian influenza surveillance system. We thus normalized values by the average value within each of those periods (1994-2003, 2004-2008 and 2009-2018) to arrive at a normalized peak number of ILI cases per 1000 consultations in Australia.

### Historical frequencies of influenza B lineages

To estimate historical frequencies of B/Victoria and B/Yamagata, we downloaded data on lineage and date and country of isolation for all influenza B isolates on the Global Initiative on Sharing All Influenza Data (GISAID) website collected until 12/31/2019. To complement these data, we searched the NCBI Influenza Virus Database for all protein-coding HA sequences of influenza B viruses isolated from humans and excluding laboratory strains (information on passage history for GISAID entries was scarce and non-standardized and so we did not filter out laboratory strains from the GISAID data). Because lineage information was missing for virtually all sequences retrieved from NCBI, we used BLAST to assign each sequence to either B/Victoria or B/Yamagata based on the highest bit score match with reference sequences B/Victoria/2/87 and B/Yamagata/16/1988.

We combined data from both databases to estimate the frequency of B/Yamagata and B/Victoria isolates in each season. Isolates collected in year *y* in Europe or North America were assigned to season *y* − 1/*y*, if collected before October, and to season *y*/*y* + 1, if collected in October-December. Because most European and North American isolates were collected before October of the respective year (median across years = 83% for Gisaid and 82% for NCBI), we assumed isolates with missing month of collection in those regions were collected before October (and thus belonged to the season beginning in the previous calendar year).

Isolates with the same name but reported for different countries or seasons were considered separately. We condensed multiple occurrences of the same isolate in the same country and season (within or across datasets) into one, disregarding isolates for which different lineages were assigned in different countries/seasons. Using isolates present in both databases, we found that our BLAST lineage assignment matched the lineage reported on GISAID in 98% (3,159/3,217) of cases. We disregarded isolates for which our BLAST assignment and the reported GISAID assignment disagreed. The final dataset consisted of 35,158 isolates, 23 of which (0.07%) were represented more than once (in different countries or seasons). We estimated the frequency of a lineage as the number of isolates belonging to that lineage divided by the total number of influenza B isolates collected in a season.

Because all 23 isolates collected in New Zealand and Australia in the 1990s were B/Yamagata (Figure S2), and because previous work suggests B/Victoria circulated only in East Asia during that period (Shaw et al., 2002), we assumed the frequency of B/Yamagata in New Zealand and Australia to be 100% from 1988 to 2000, even though no isolates were available for several individual years within that range. In 2001 and 2003, when both lineages are known to have been circulating in New Zealand and Australia but fewer than 10 isolates were available for the two countries combined in the sequence databases, we used frequencies estimated from all other countries combined. Frequencies estimated from all other countries combined were strongly correlated with estimates based on isolates from Australia and New Zealand only (Pearson’s correlation coefficient = 0.88, 95% CI 0.76-0.95). We also considered using frequencies estimated from isolates collected in the United States, which were also correlated with frequencies in Australia and New Zealand (Figure S2), but the correlation was weaker (0.69, 95% CI 0.42-0.85). For the sensitivity analysis of lineage frequencies in the 1990s, we re-fitted the model using frequencies from all countries combined for all years from 1988 on in which fewer than 10 isolates were available for New Zealand and Australia combined, including years in the 1990s.

We hoped to use the sequence databases to get more reliable estimates of lineage frequencies in the 1980s than those provided by early antigenic characterization (Rota et al., 1990), but fewer than 10 isolates were available for each year before 1988. To accommodate uncertainty, we grouped infections with B/Victoria and B/Yamagata before 1988 with infections by the ancestral influenza B strains circulating before the lineages split.

We compared our estimates of lineage frequencies based on sequence data to estimates based on antigenic characterization of circulating strains from epidemiological surveillance reports (Figure S2). Surveillance reports from Australia are available from the Australian Government’s Department of Health website (Australian Government Department of Health, 2019). Surveillance reports from New Zealand are available from the website of New Zealand’s Institute of Environmental Science and Research (ESR) (Institute of Environmental Science and Research, 2020). Although reports from New Zealand are only available from 2003 on, Fig. 27 of the 2012 report shows B/Victoria and B/Yamagata frequencies from 1990 to 2002 (without reporting the number of isolates used to estimate those frequencies). Annual summaries of influenza surveillance in the United States are published by the Centers for Disease Control and Prevention (e.g., Garten et al. (2018)).

### Age distributions of cases in non-surveillance data

We applied the model to the age distributions of influenza B isolates available on GISAID. In these analyses, isolate data are used in two different ways: both to estimate historical lineage frequencies and to estimate the age distributions of cases to which the model is fitted. While historical lineage frequencies are calculated based on all GISAID isolates with lineage information (and complemented with isolates from the NCBI Influenza Virus Database; see above), only a subset of GISAID isolates are annotated with both the isolate’s lineage and the host’s age, and therefore only a subset can be used to estimate the age distributions of cases. We analyzed the age distributions of isolates from the European Union and China because those regions have many isolates with both lineage and age information (4,702 isolates from 2006-2018 for the European Union, 1,880 isolates from 2005-2019 for China) and easily available demographic data for the general population (we excluded the United States because historically high influenza vaccination coverage would require incorporating vaccination into the model.) To increase statistical power for the estimates of historical lineage frequencies, we included isolates from European countries outside the European Union and countries in East Asia besides China (South Korea, Japan, Mongolia and Taiwan) represented in GISAID (Figure S2). Individual countries are represented in different proportions in the lineage frequency and age distribution datasets (Figure S24). Thus, if lineage frequencies vary in space within Europe or within East Asia, our estimates of infection history probabilities might be biased relative to the true infection histories associated with the age distributions used to fit the model.

Since B/Yamagata was the only lineage circulating in Europe in the mid to late 1990s, when isolate data were more abundant, we assumed B/Yamagata was completely dominant throughout the entire 1990s, as we did for New Zealand. We relaxed this assumption by letting B/Victoria be the only circulating lineage in 1988-1993 in Europe, consistent with its high frequency in the scarce isolate data from that period.

For both the EU and China, we assumed constant intensity scores across seasons (*S*_*i*_ = 1) and used the cumulative incidence of cases within seasons (*ϕ*_*i*_(*b, w*)) from New Zealand to model infection in the first year of life.

### Demographic data

We obtained annual age distributions for the general population by single year of age from governmental websites from New Zealand (StatsNZ, 2019), Australia (Australian Bureau of Statistics, 2019), China (National Bureau of Statistics of China, 2021) and the European Union (Eurostat, 2021).

### Model validation with independent serological data

We compared the fraction of children predicted by our model to have been previously infected with B/Victoria, B/Yamagata or either lineage with the fraction of children that had detectable antibodies against the corresponding lineage (or any influenza B strain) in the Netherlands (Bodewes et al., 2011). Sera from children 0-7 years old collected between February 2006 and June 2007 were tested using the hemagglutination inhibition assay against a panel of reference B/Victoria and B/Yamagata strains as well of strains isolated in the Netherlands during the study period. Sera were considered positive if their hemagglutination inhibition titer was ≥ 10 against at least one strain from the corresponding set (all influenza B strains, B/Victoria strains, and B/Yamagata strains). We compared these data with predictions under the maximum likelihood-parameter estimates of our model fitted to the complete New Zealand data.

We also used the seroprevalence data to independently estimate the annual probability of infection for preschoolers and school-age children, equivalent to the *β*_1_ and *β*_2_ parameters in our model. Assuming a constant instantaneous attack rate *α*, an individual of age *A* years is still naive (and therefore seronegative) to influenza B with probability *P*_*N*_ (*A*) given by:

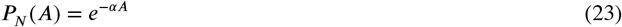

The probability of observing *X* seronegative individuals in a sample of *n* individuals of age *A* can be calculated assuming *X* ∼ Binomial[*n, P*_*N*_ (*A*)], and *α* thus can be estimated by maximum likelihood. The annual attack rate can then be calculated from the instantaneous attack rate *α* as *β*_Netherlands_ = 1 − *e*^−*α*^.

We make two modifications to Eq. (23) to account for the presence of maternal antibodies early in life and for uncertainty in the age of individuals when their serum was collected. First, we assume individuals spend a time *m* (in units of years) fully protected against influenza B due to the presence of maternal antibodies. Consistent with the fraction of children under the age of 1 with detectable antibodies to influenza B (Bodewes et al., 2011), we assumed *m* = 0.5 year. Second, because ages were reported at the resolution of one year (e.g. an individual 2.6 years old is reported as being 2 years old), we assume individuals with recorded age *A* were sampled at a randomly distributed time *T* ∈ [0, 1) during the interval between the ages of *A* and *A* + 1. Thus, we let *P*_*N*_ (*A*) be given by the expectation over *T*:

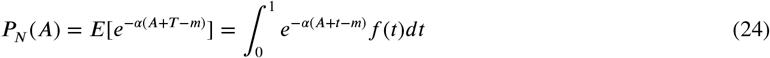

where *f* (*t*) is the probability density function of *T*. Assuming *T* is uniformly distributed between 0 and 1 (i.e., *f* (*t*) = 1), we have:

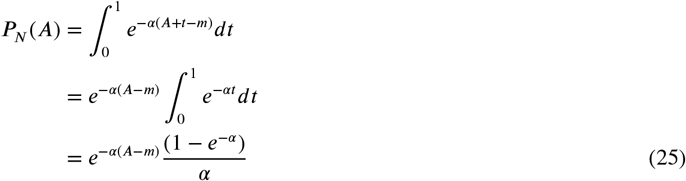

valid for *A > m* and *α >* 0. Letting *α*_1_ and *α*_2_ be the instantaneous attack rates for preschoolers and school-age children:

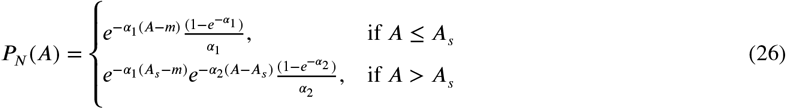

where *A*_*s*_ is the age at which children start going to school (4 years old in the Netherlands; Government of the Netherlands (2020)). Note that for school-age children (the equation for *A > A*_*s*_ on the bottom) the correction term for uncertainty in sampling is not necessary for the time spent in preschool (assumed to be exactly *A*_*s*_ years), only for the time after preschool (*A* − *A*_*s*_).

### Handling cases with missing lineage information

We assumed cases with missing lineage information in 2002 (*n* = 61), 2011 (*n* = 312) and 2019 (*n* = 206) belonged to B/Victoria, since 99% or more of identified cases in those seasons were B/Victoria (86/87 cases in 2002, 276/280 cases in 2011, 552/552 cases in 2019) as were 94%, 92% and 92% of isolates from sequence databases (for Australia and New Zealand combined). We assumed cases with missing lineage information belonged to B/Yamagata in 2013 (*n* = 37), 2014 (*n* = 77) and 2017 (*n* = 87), when the majority of identified cases were B/Yamagata (268/272, 131/138 and 473/489, respectively), as were 99%, 94% and 84% of isolates in sequence databases. Unidentified cases in other seasons were disregarded because both lineages were present at higher frequencies among identified cases. Removing unidentified cases altogether in all seasons led to similar parameter estimates.

### Sequence divergence analysis

To estimate the amount of evolution within and between lineages, we analyzed all complete HA and NA sequences from human influenza B isolates available on GISAID in July 2019. The set of isolates used in this analysis differs from the set used to estimate lineage frequencies because we required isolates to have complete sequences (although not all sequences listed as complete on GISAID were in fact complete). Two isolates collected in 2000 (B/Hong Kong/548/2000 and B/Victoria/504/2000) were deposited as B/Victoria but our BLAST assignment indicated they were in fact B/Yamagata (their low divergence from B/Yamagata strains was a clear outlier). NA sequences from isolates B/Kanagawa/73 and B/Ann Arbor/1994 were only small fragments (99 and 100 amino acids long) poorly aligned with other sequences and were thus excluded. We also excluded NA sequences from B/Yamagata isolates B/Catalonia/NSVH100773835/2018 and B/Catalonia/NSVH100750997/2018 because they were extremely diverged (60% and 38%) from the reference strain B/Yamagata/16/88 and aligned poorly with other sequences.

To compare sequence diversity within and between lineages over time, we aligned sequences using MAFFT v. 7.310 (Katoh et al., 2002) and calculated percent amino acid differences in pairs of sequences from the same lineage and in pairs with one sequence from each. For each year, we sampled 100 sequences from each lineage (or used all sequences if 100 or fewer were available) to limit the number of pairwise calculations. To estimate how much B/Yamagata and B/Victoria evolved since the late 1980s, we calculated percent amino acid differences between each B/Yamagata and B/Victoria sequence and the corresponding HA and NA sequences of reference strains B/Yamagata/16/88 and B/Victoria/2/87. Unlike in the analysis of pairwise divergence within each time point, we used all sequences from each lineage in each year. We excluded sites in which one or both sequences had gaps or ambiguous amino acids.

To compare HA and NA divergence between influenza B lineages with divergence between influenza A subtypes, we downloaded complete HA and NA sequences from H3N2 and H1N1 isolated since 1977 and available on GISAID in August 2019. Homologous sites in the HA of H3N2 and H1N1 are difficult to identify by conventional sequence alignment, and instead we used the algorithm by Burke & Smith (Burke and Smith, 2014) implemented on the Influenza Research Database website (Influenza Research Database, 2019). Both H3N2 and H1N1 sequences were aligned with the reference H3N2 sequence A/Aichi/2/68. We verified that this method matched sites on the stalk and head of the H1N1 HA with sites on the stalk and head of H3N2 HA by comparing the resulting alignment with the alignment in Fig. S2 of Kirkpatrick et al. (2018). To limit the total number of influenza A sequences analyzed we randomly selected 100 H3N2 and 100 H1N1 sequences for years in which more than 100 sequences were available and used all available sequences for the remaining years. Isolates A/Canterbury/58/2000, A/Canterbury/87/2000 and A/Canterbury/55/2000 were excluded because both H1N1-like and H3N2-like sequences were available under the same isolate name on GISAID.

## Data Availability

Data in the final format used in the analyses are available at https://github.com/cobeylab/influenza_B_distributions

## Data availability

The data used in the analyses are available at https://github.com/cobeylab/influenza_B_distributions.

## Code availability

Code implementing the analyses and figures is available at https://github.com/cobeylab/influenza_B_distributions.

## Acknowledgments

Frank Wen helped with data collection from GISAID. Three anonymous reviewers provided helpful comments and suggestions. This work was completed in part with resources provided by the University of Chicago Research Computing Center. This project has been funded in part with Federal funds from the National Institute of Allergy and Infectious Diseases, National Institutes of Health, Department of Health and Human Services under grant DP2 AI117921 and CEIRS Contract No. HHSN272201400005C awarded to S.C. M.V. was supported by a William Rainey Harper Dissertation Fellowship by the University of Chicago. K.K. was supported by MIDAS CIDID Center of Excellence (U54-GM111274), V.D. was supported by NIH CEIRS Contract No. HHSN272201400006C, C.D. was supported by an Early Career Fellowship (1113269) from the Australian National Health and Medical Research Council. The content is solely the responsibility of the authors and does not necessarily represent the official views of the funding sources.

## Author contributions

M.C.V., S.C., C.D., K.K., and V.D. designed the study. M.C.V., S.C., P.A. and K.K. developed the mathematical models. M.C.V. wrote the code and performed the analysis. G.F.R. provided data on influenza B seroprevalence in children. L.L., T.W. and Q.S.H. contributed influenza B case data. M.C.V. wrote the first draft of the manuscript, and all authors approved the final version.

## Competing interests

No competing interests declared.

## Supplementary information

**Table S1.**
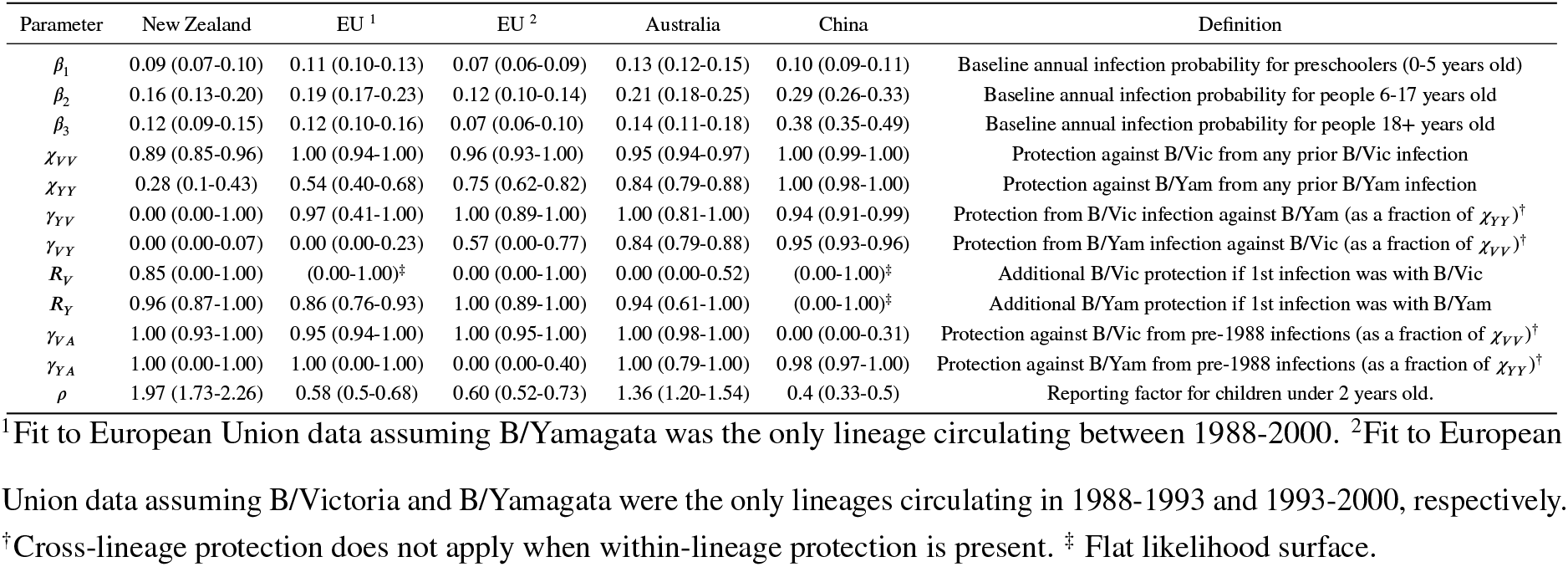
Parameter estimates from the New Zealand data compared to estimates based on additional data sets.

**Figure S1.**
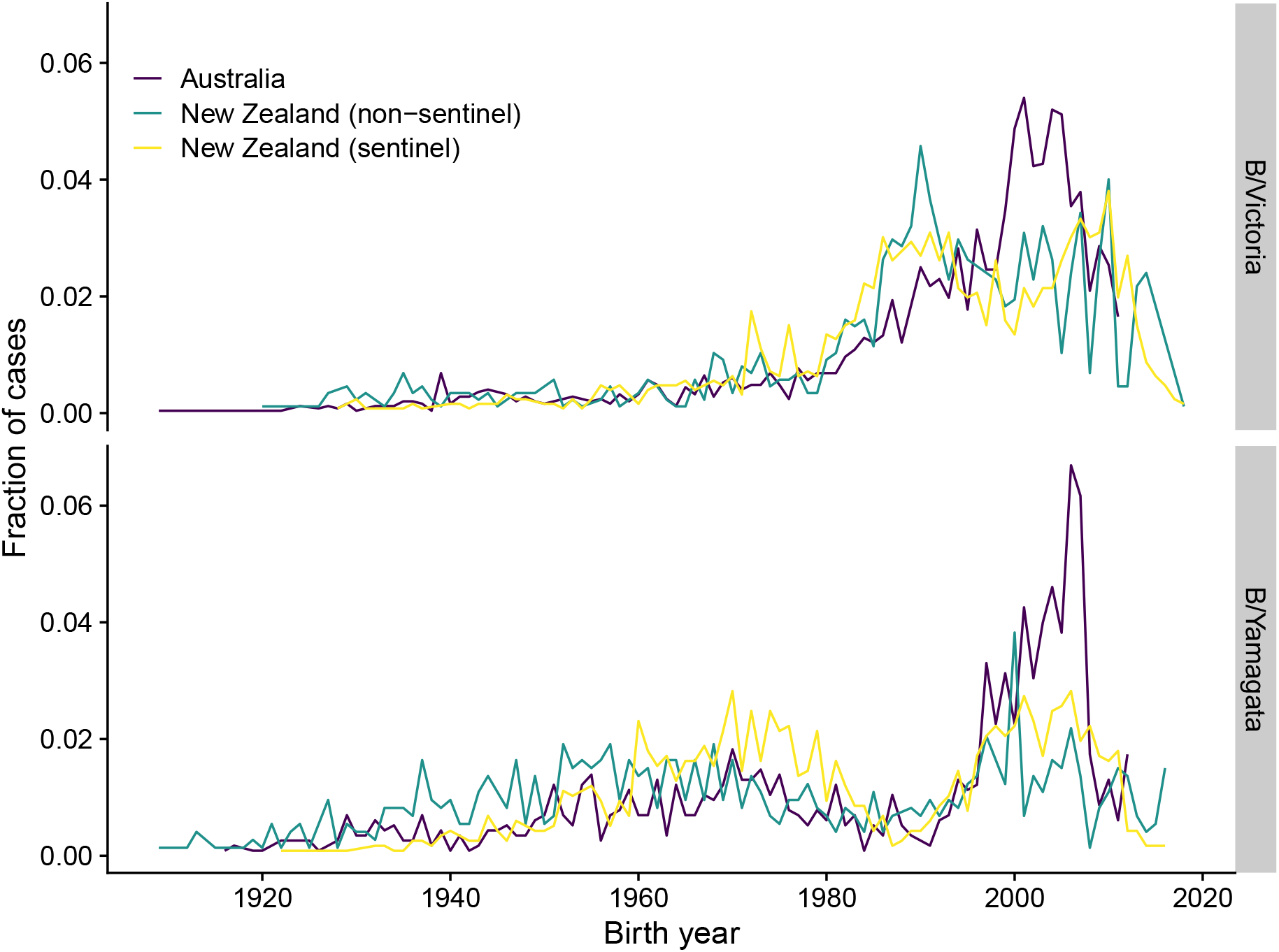
Distribution of medically attended influenza B cases in New Zealand (2001-2019) and Australia (2002-2013) by birth year of the patient. The fraction of cases in each birth year was calculated relative to all cases observed for each lineage (separately by type of surveillance in New Zealand).

**Figure S2.**
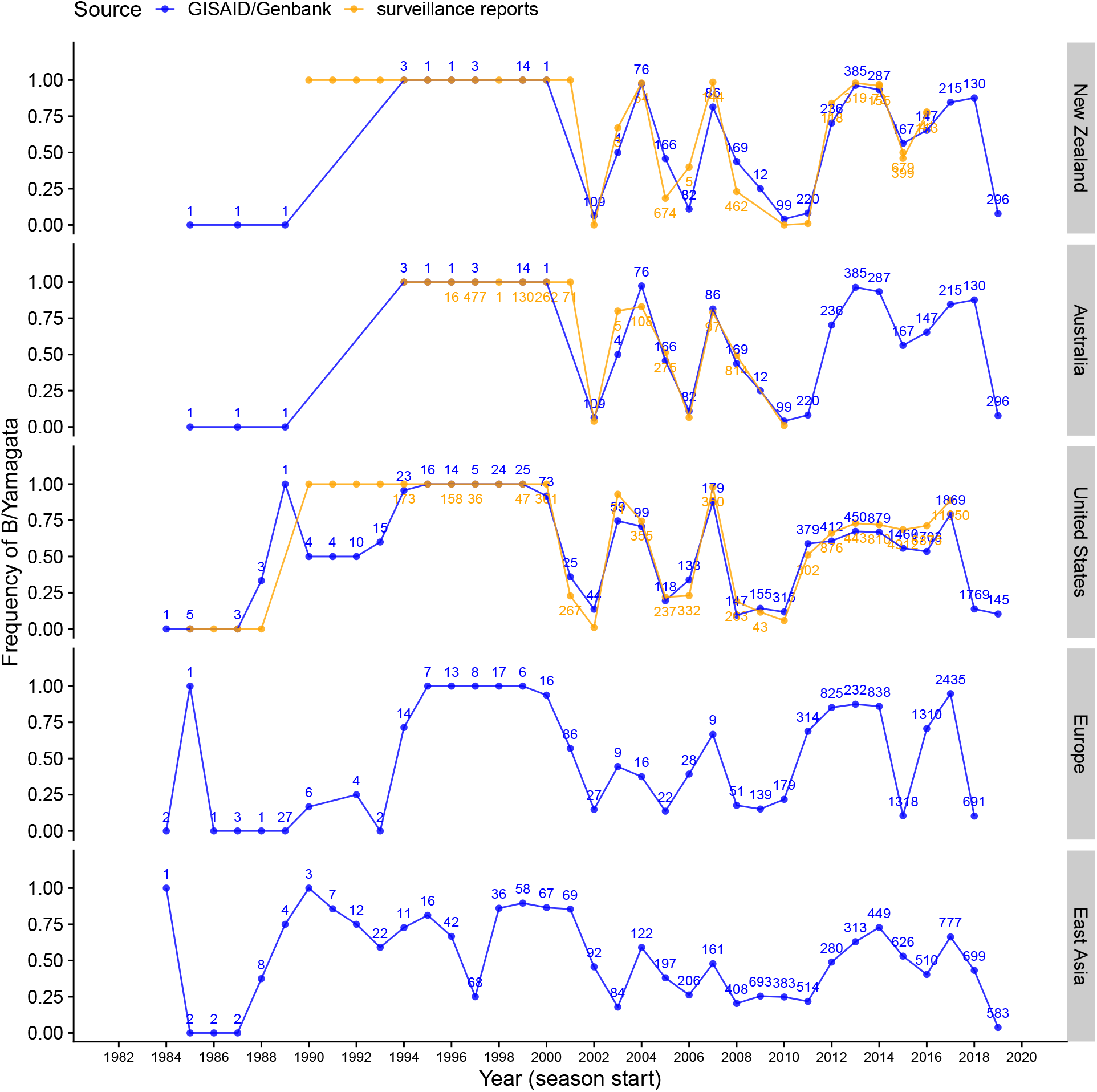
Comparison of lineage frequency estimates based on sequence data and surveillance reports. Frequencies are shown by annual influenza seasons, which span multiple calendar years in the Northern hemisphere but are contained in a single calendar year in the Southern Hemisphere (e.g., 2006 refers to the 2006/2007 influenza season in the United States, Europe and East Asia and to the 2006 season in Australia and New Zealand). Numbers indicate the number of isolates collected in that season and deposited on GISAID or the NCBI Influenza Virus Database (blue) or tested by surveillance (orange; the total number of isolates tested was not reported in some of the surveillance reports). Sequence database isolates from Australia and New Zealand were grouped together to estimate lineage frequencies.

**Figure S3.**
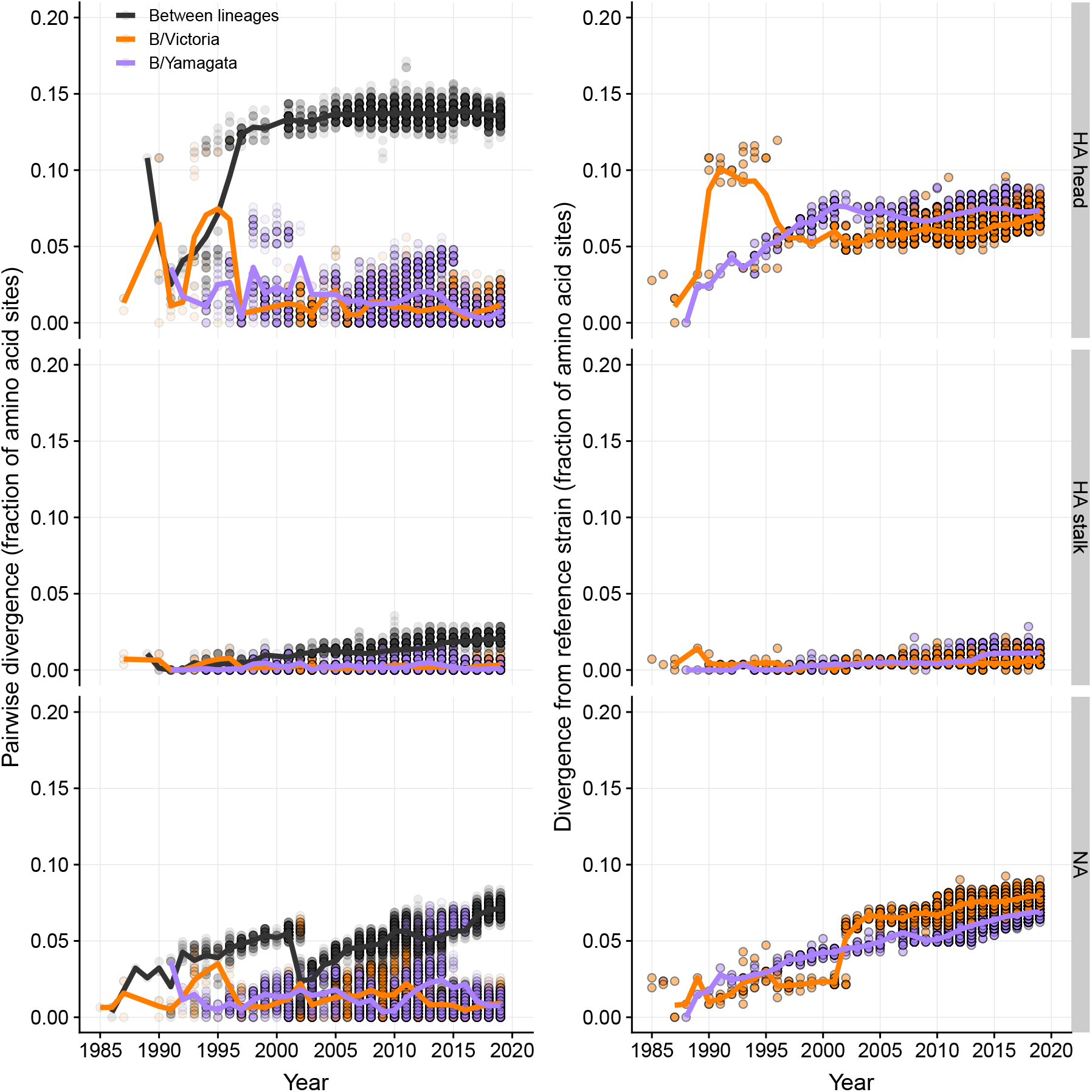
Amino acid divergence in the hemagglutinin (HA) and neuraminidase (NA) proteins within and between influenza B lineages. The left panel shows the fraction of amino acid sites that differ between pairs of sequences in each season (each point is a pair). Up to 500 randomly chosen pairs are shown, while the solid lines represent the annual average calculated from all pairs given two samples of up to 100 sequences from each lineage (i.e, up to 4,950 pairs). The right panel shows divergence of strains circulating at different times from reference strains B/Victoria/2/87 and B/Yamagata/16/88.

**Figure S4.**
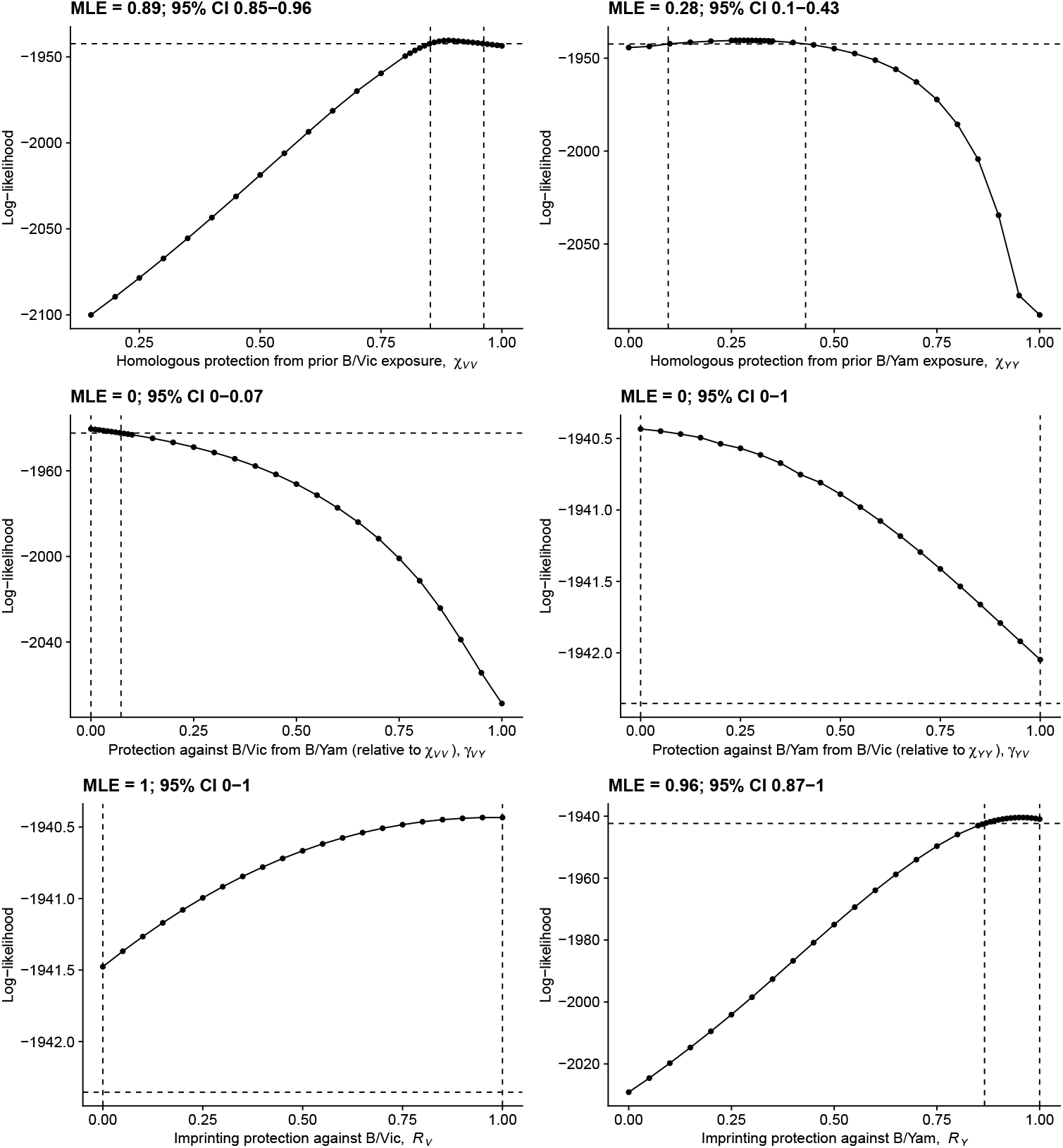
Likelihood profiles for protection parameters estimated from the complete New Zealand data. The 95% confidence interval based on a likelihood ratio test with one degree of freedom is indicated by the vertical dashed lines.

**Figure S5.**
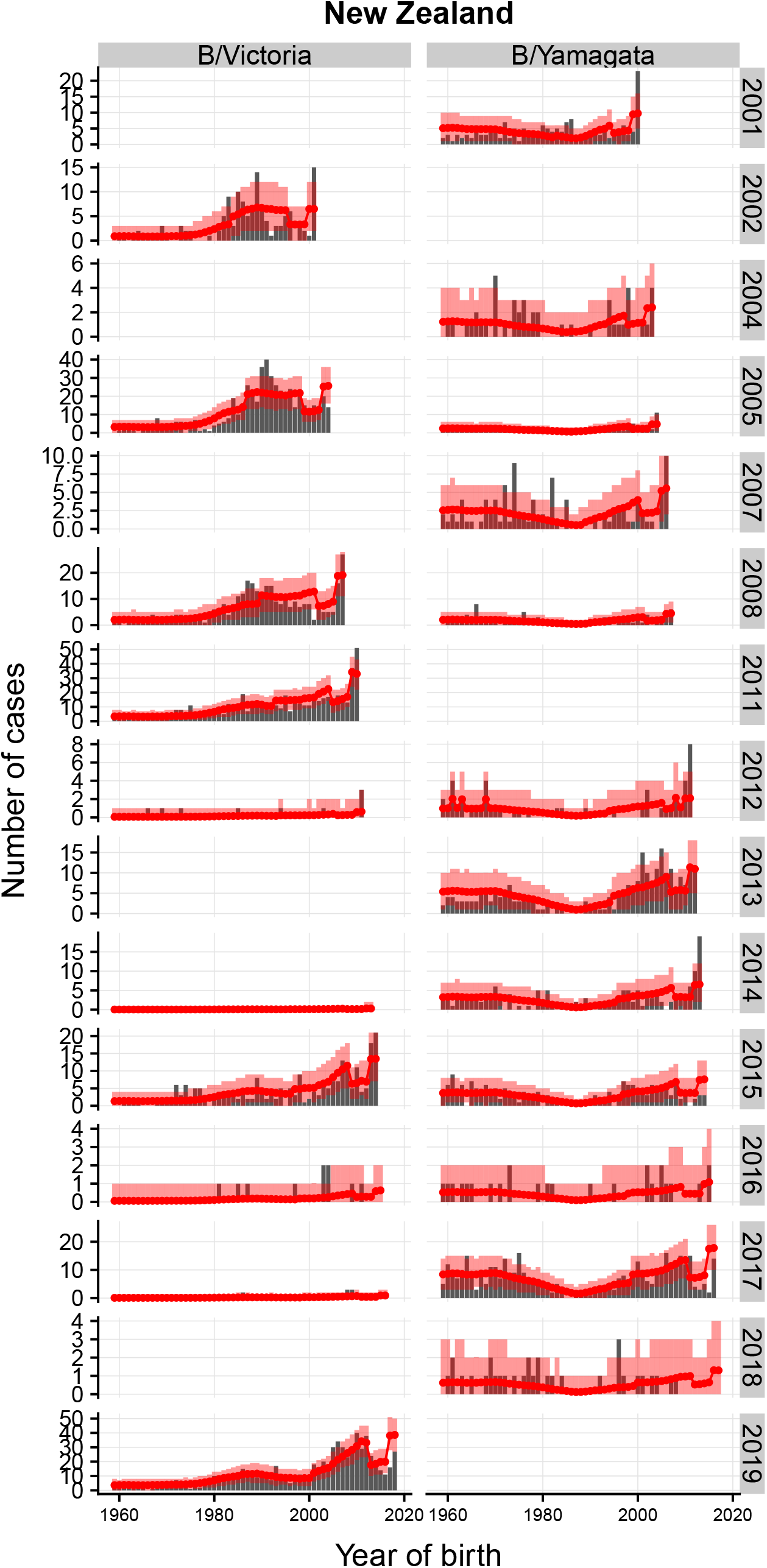
Observed and predicted distributions of influenza B cases in New Zealand by observation year. Vertical red bars are 95% bootstrap confidence intervals. Season/lineage combinations with fewer than 5 cases were omitted to improve visualization.

**Figure S6.**
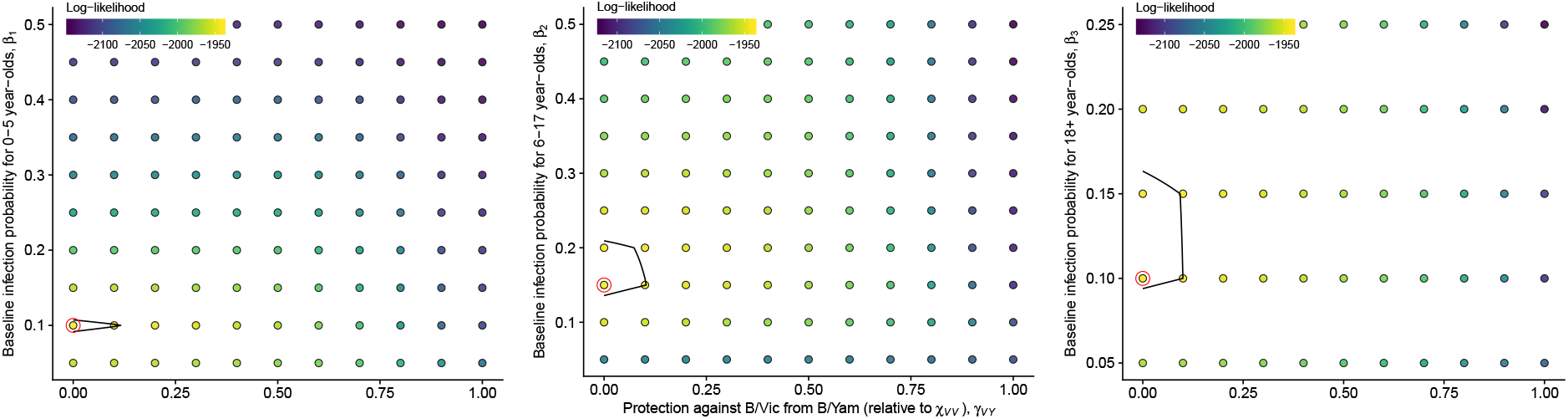
Bivariate profiles of age-specific baseline probabilities of infection and protection from B/Yamagata against B/Victoria. The red circle indicates the maximum likelihood value in the profile. The contour indicates the 95% confidence interval based on a bilinear interpolation of the profiled points.

**Figure S7.**
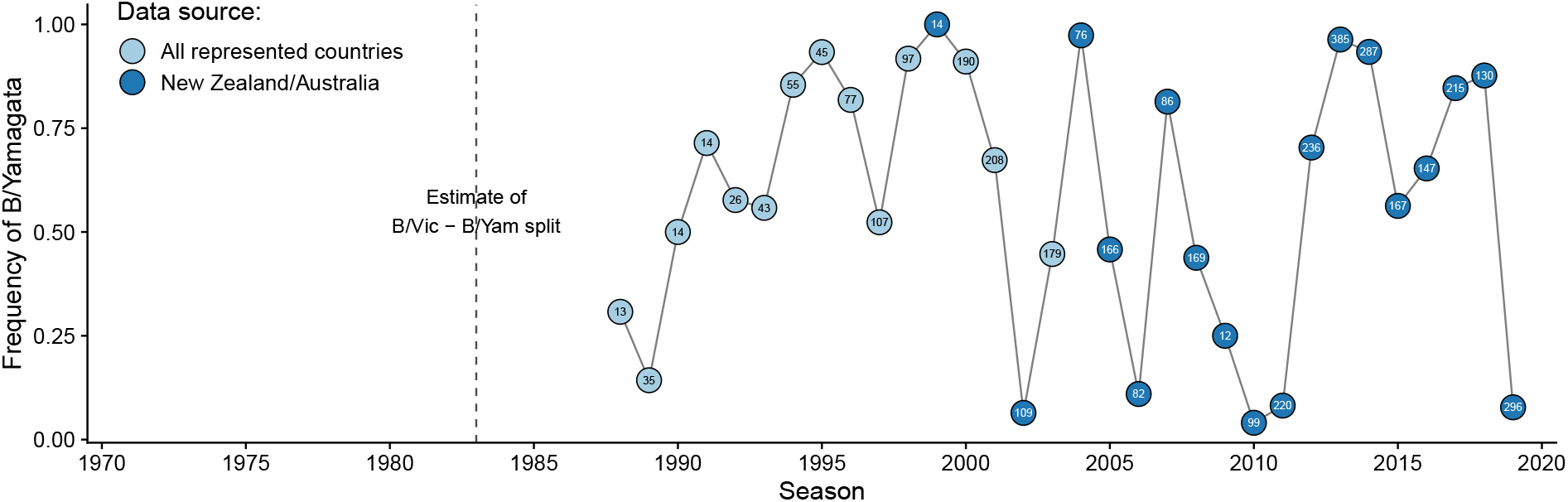
Lineage frequencies used in sensitivity analysis to test the effect of having only B/Yamagata in New Zealand in the 1990s. If fewer than 10 isolates were reported in New Zealand and Australia combined in a season, we estimated frequencies using isolates from all countries represented in the databases. The number inside each circle indicates the number of isolates used to estimate lineage frequencies in each season.

**Figure S8.**
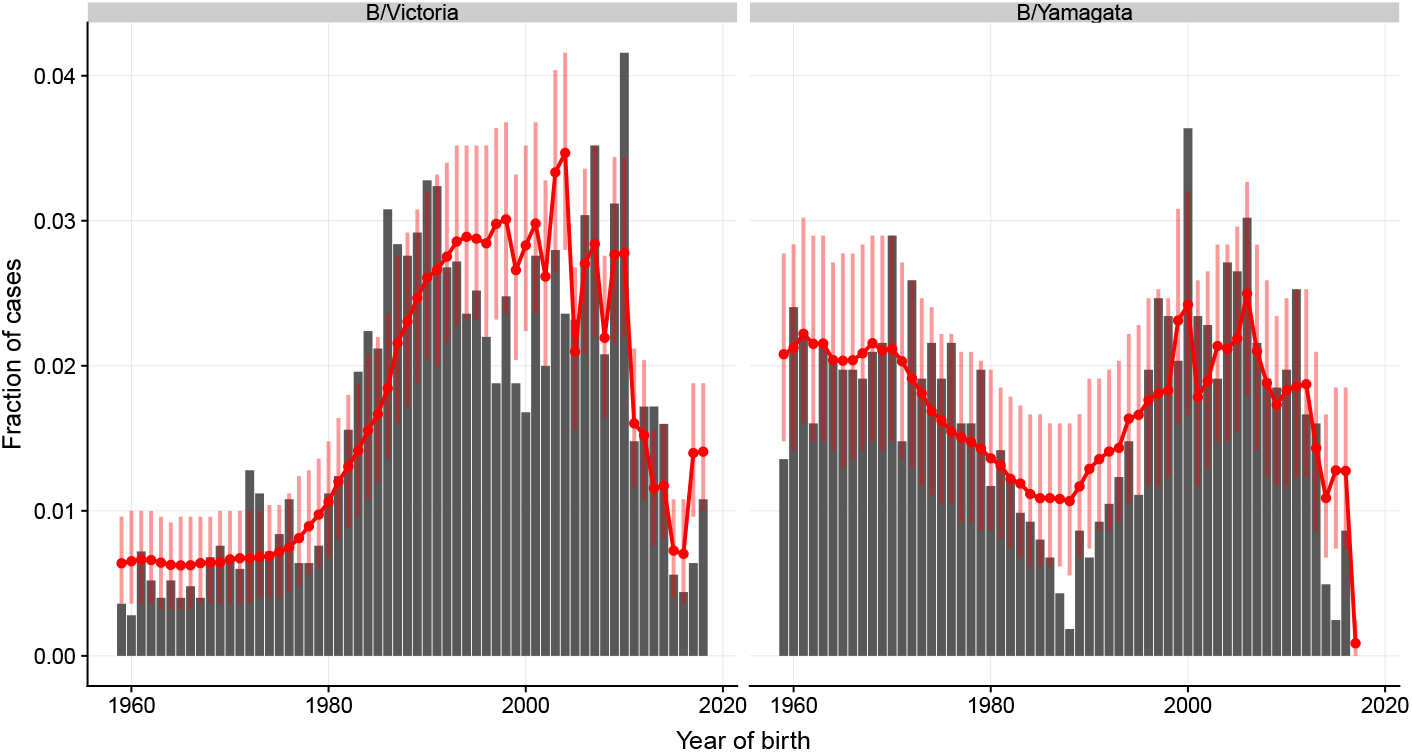
Model predictions from the sensitivity analysis of the frequency of B/Yamagata in New Zealand in the 1990s. Red lines and dots show the predicted distribution under the model. Vertical bars are 95% bootstrap confidence intervals.

**Figure S9.**
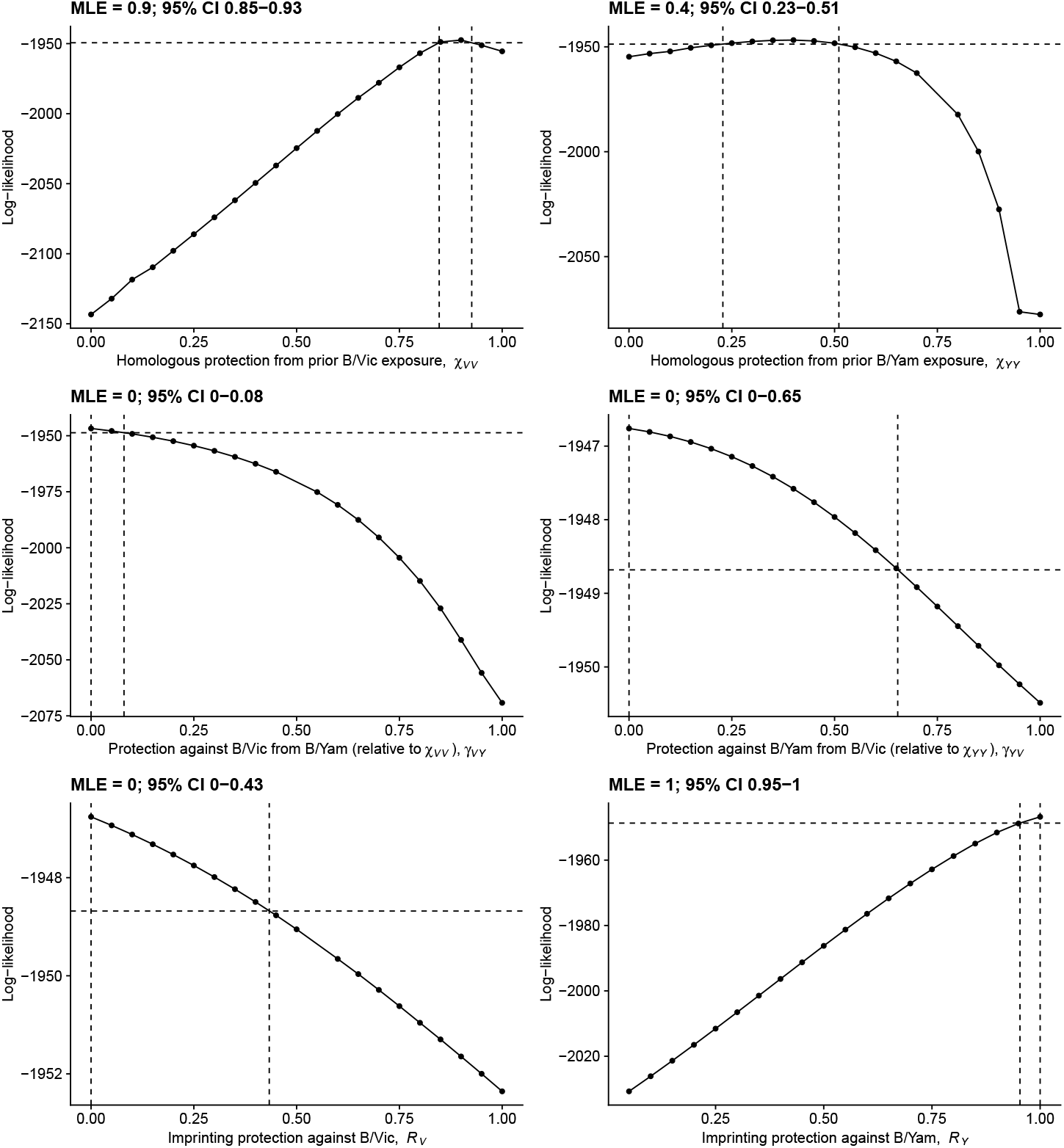
Likelihood profiles for protection parameters estimated assuming B/Victoria was the only lineage circulating between 1983 and 1990. The 95% confidence interval based on a likelihood ratio test with one degree of freedom is indicated by the vertical dashed lines.

**Figure S10.**
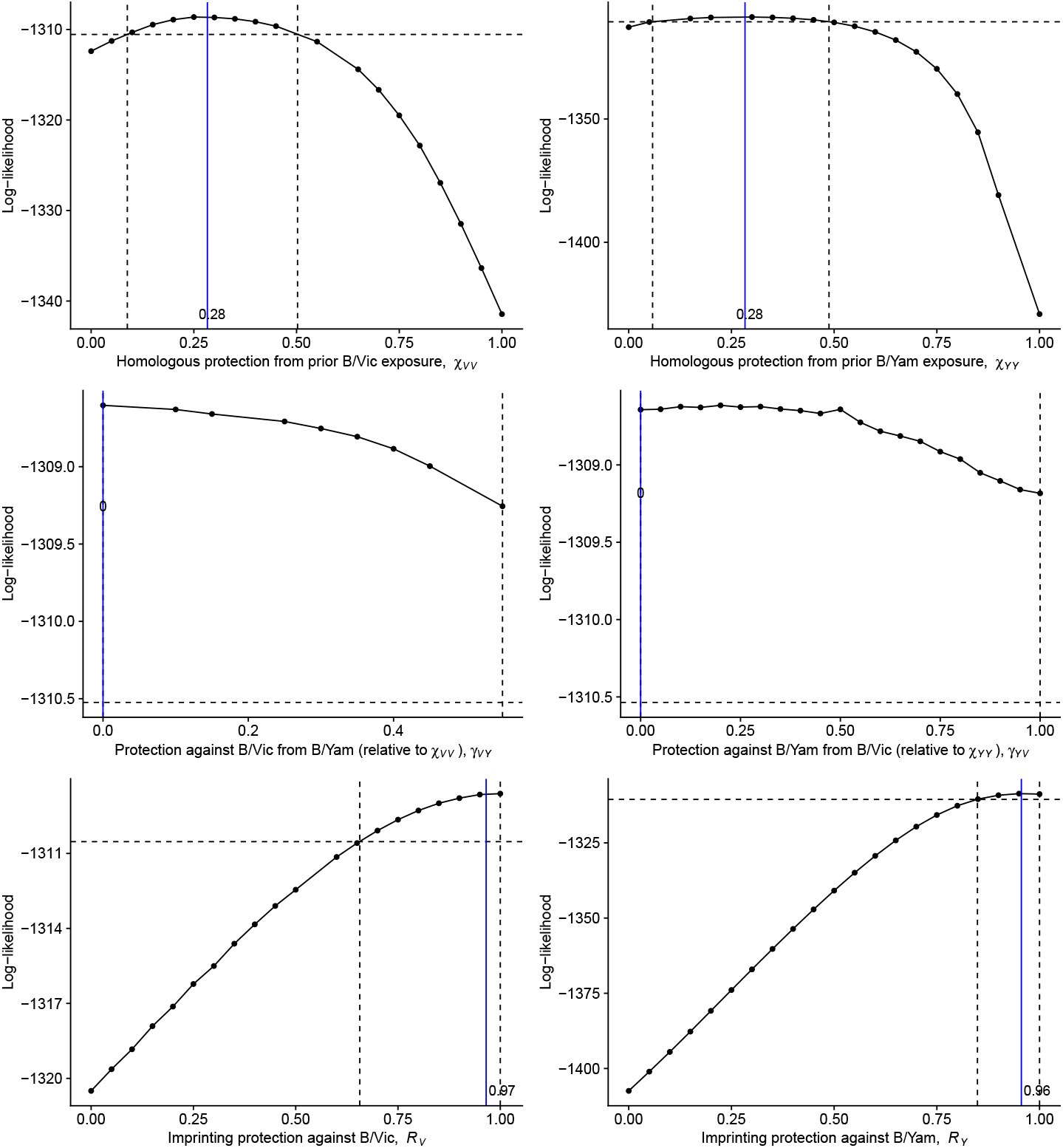
Likelihood profiles for protection parameters estimated from data simulated under strong imprinting protection but moderate within-lineage protection against B/Victoria. Parameter values used for the simulation are represented by the vertical blue lines and the numbers next to them. The other parameters beside *R*_*V*_ and *χ*_*VV*_ were set to their maximum likelihood values from the main analysis. The 95% confidence interval based on a likelihood ratio test with one degree of freedom is indicated by the vertical dashed lines.

**Figure S11.**
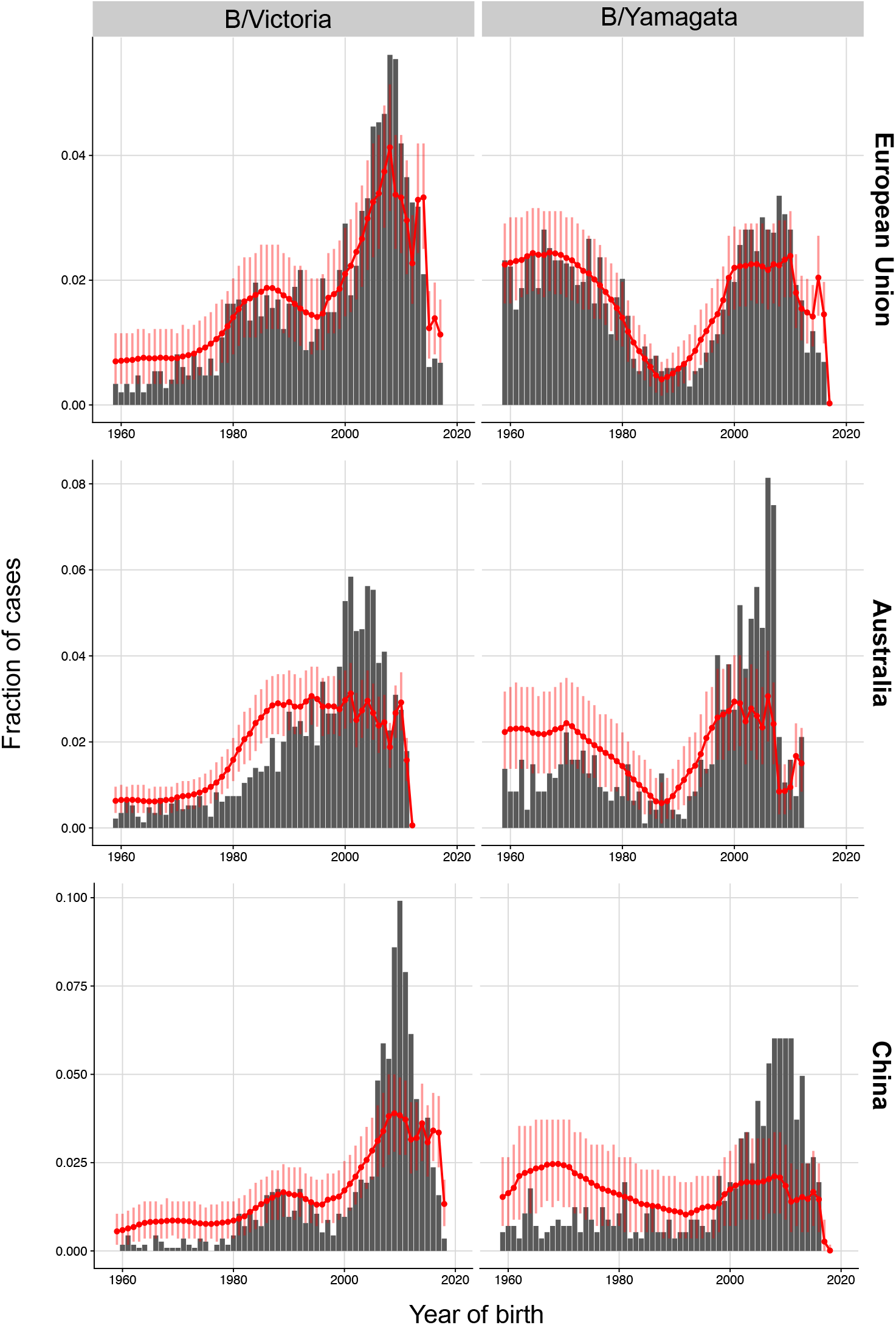
Model predictions for additional data sets using protection parameters estimated from the New Zealand data. Red lines and dots show the predicted distribution under the model. Vertical bars are 95% bootstrap confidence intervals.

**Figure S12.**
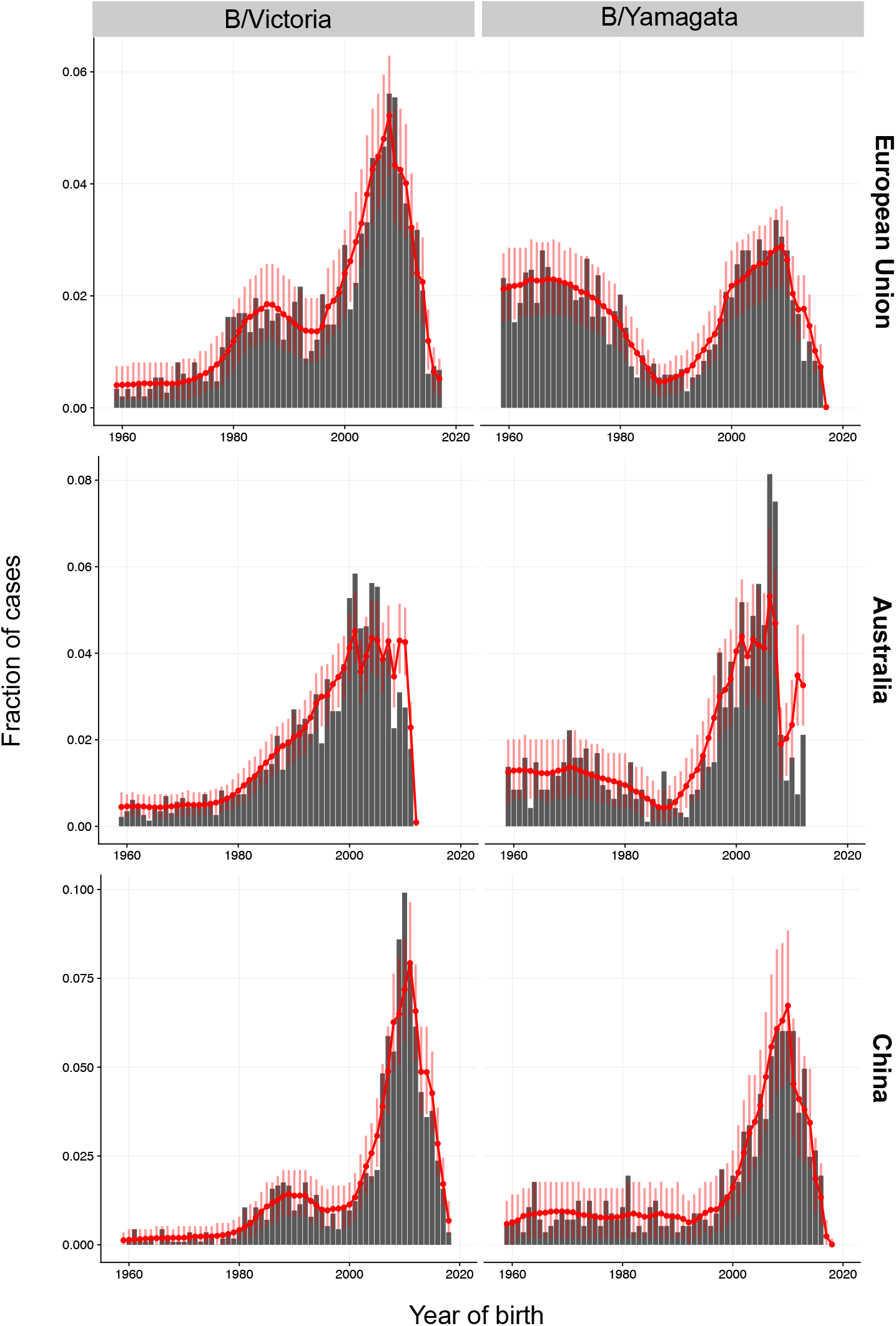
Model predictions for additional data sets using parameters re-estimated for each data set. Red lines and dots show the predicted distribution under the model. Vertical bars are 95% bootstrap confidence intervals.

**Figure S13.**
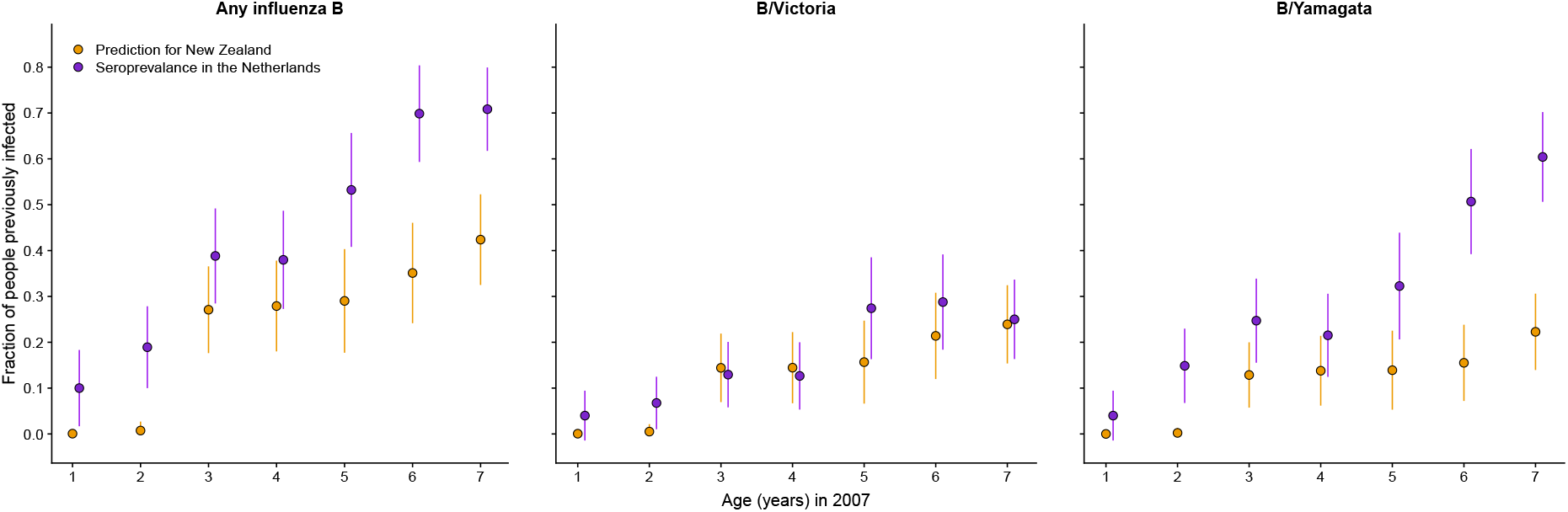
Frequency of past influenza B infections in children predicted by the model compared with observed seroprevalence in the Netherlands. The fraction of children previously infected with influenza B in the Netherlands in 2006-2007 was estimated by Bodewes et al. (2011) as the fraction having serum antibodies against at least one influenza B strain from a panel of viruses (left panel), at least one B/Victoria strain (center panel) or at least one B/Yamagata strain (right panel). Bars show 95% binomial confidence intervals. Fractions for Australia and New Zealand were generated independently from the seroprevalence data by fitting a statistical model to medically attended influenza B infections. For the model predictions, binomial confidence intervals assume a sample size equal to the number of children with the corresponding age in the seroprelavence data.

**Figure S14.**
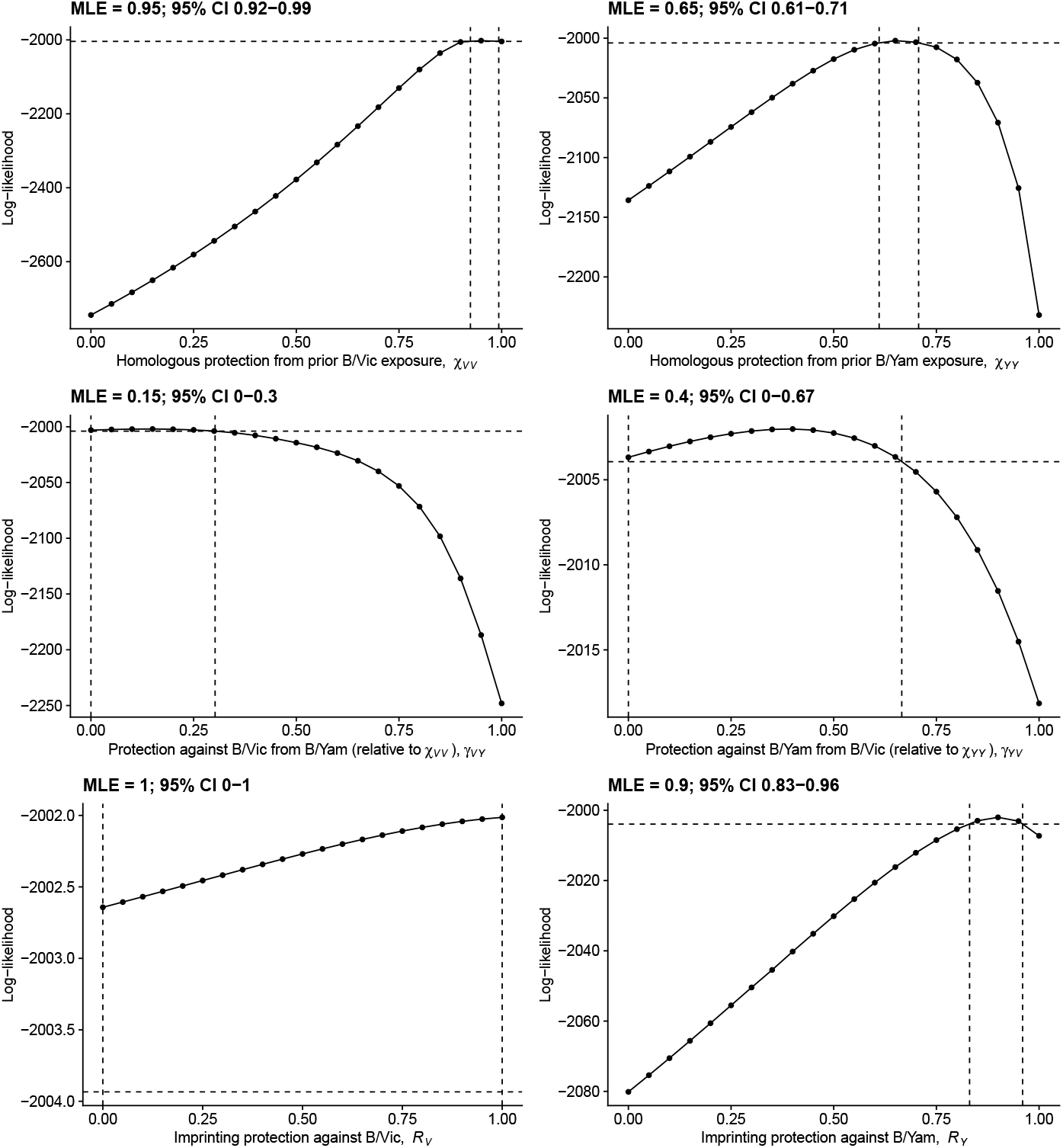
Likelihood profiles estimated under baseline annual infection probabilities constrained to values inferred from Dutch seroprevalence data. The 95% confidence interval based on a likelihood ratio test with one degree of freedom is indicated by the vertical dashed lines.

**Figure S15.**
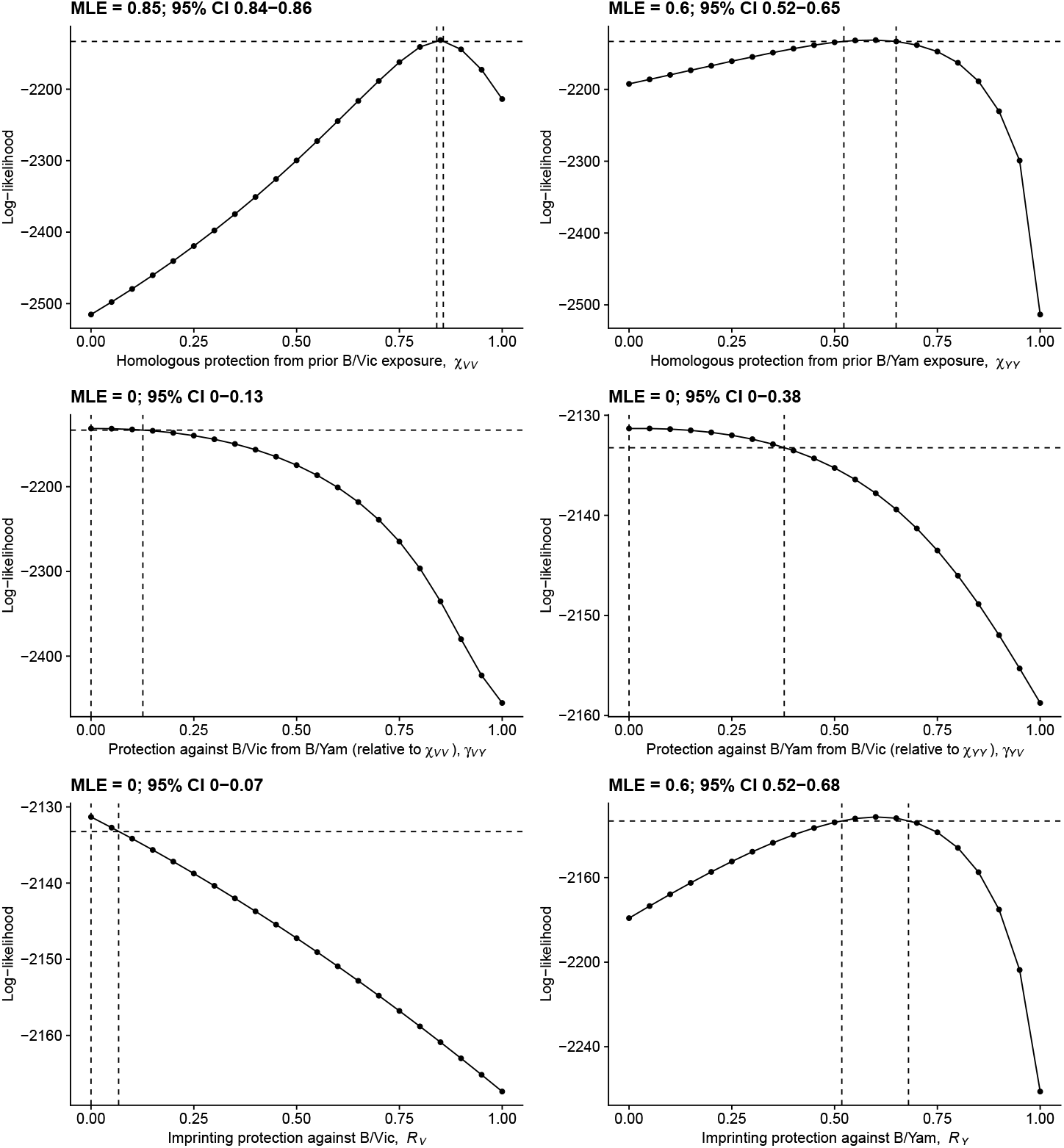
Likelihood profiles estimated under a 30% baseline annual infection probability across age groups. The 95% confidence interval based on a likelihood ratio test with one degree of freedom is indicated by the vertical dashed lines.

**Figure S16.**
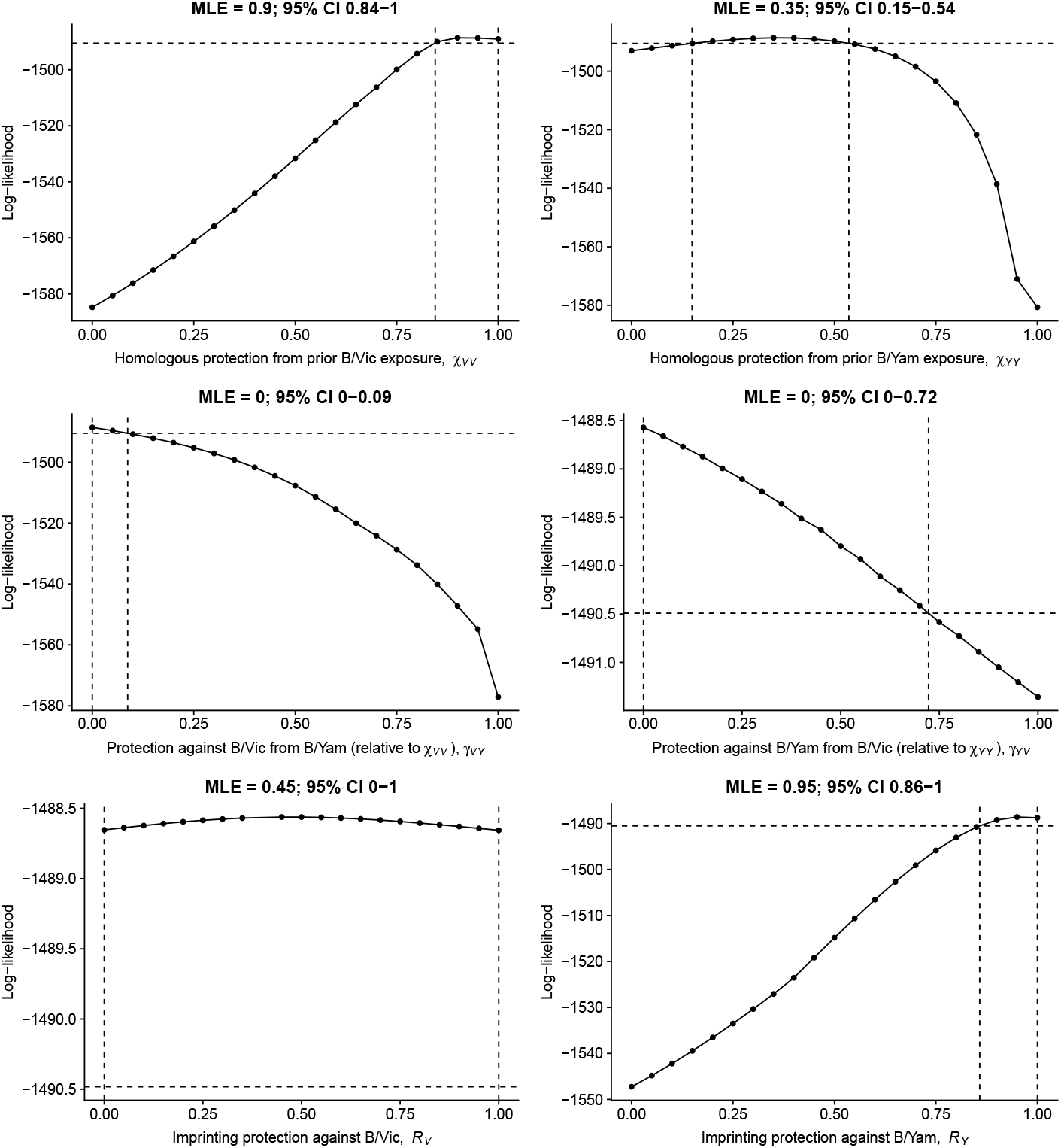
Likelihood profiles for protection parameters estimated from surveillance cases alone. The 95% confidence interval based on a likelihood ratio test with one degree of freedom is indicated by the vertical dashed lines.

**Figure S17.**
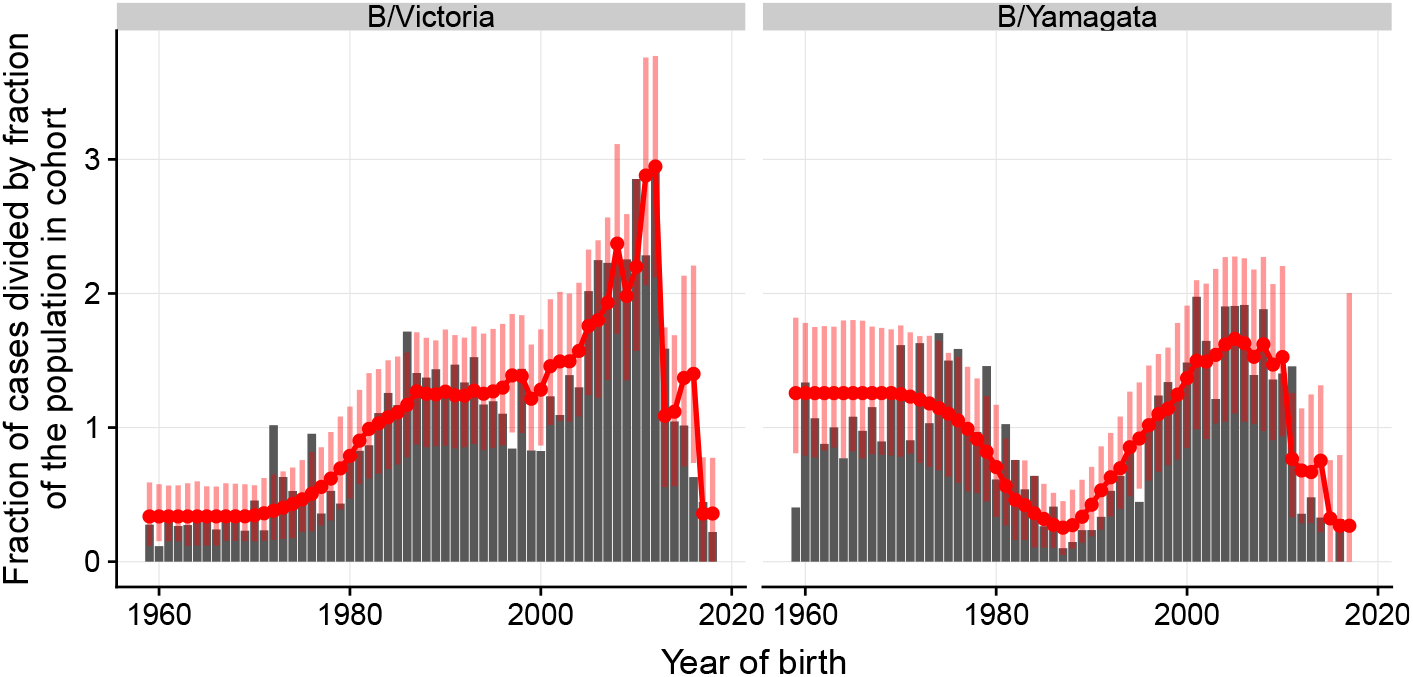
Age distributions predicted by the model fitted to surveillance cases only. Red lines and dots show the predicted distribution under the model. Vertical bars are 95% bootstrap confidence intervals.

**Figure S18.**
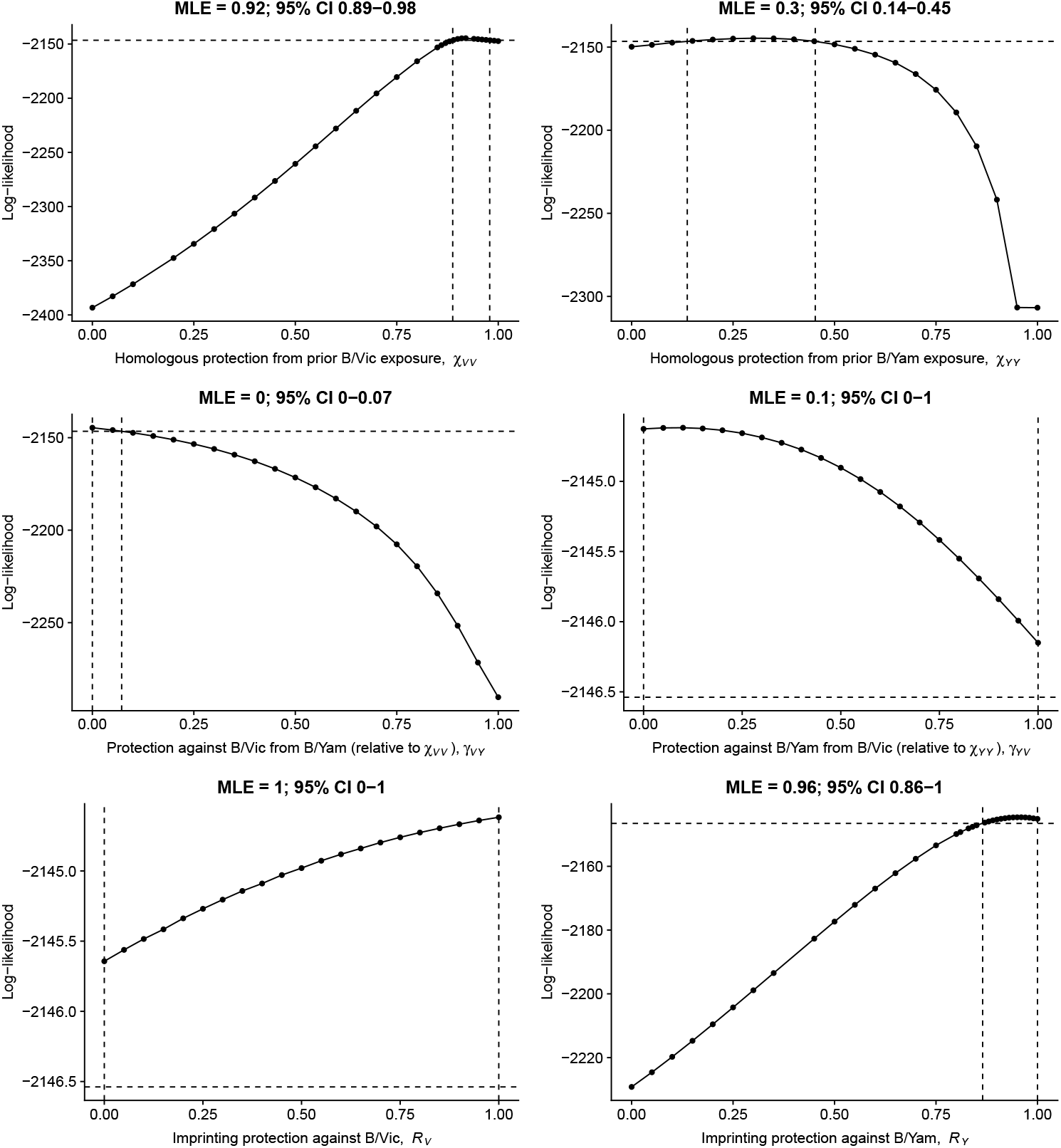
Likelihood profiles for protection parameters estimated from birth years starting in 1952 instead of 1959. The 95% confidence interval based on a likelihood ratio test with one degree of freedom is indicated by the vertical dashed lines.

**Figure S19.**
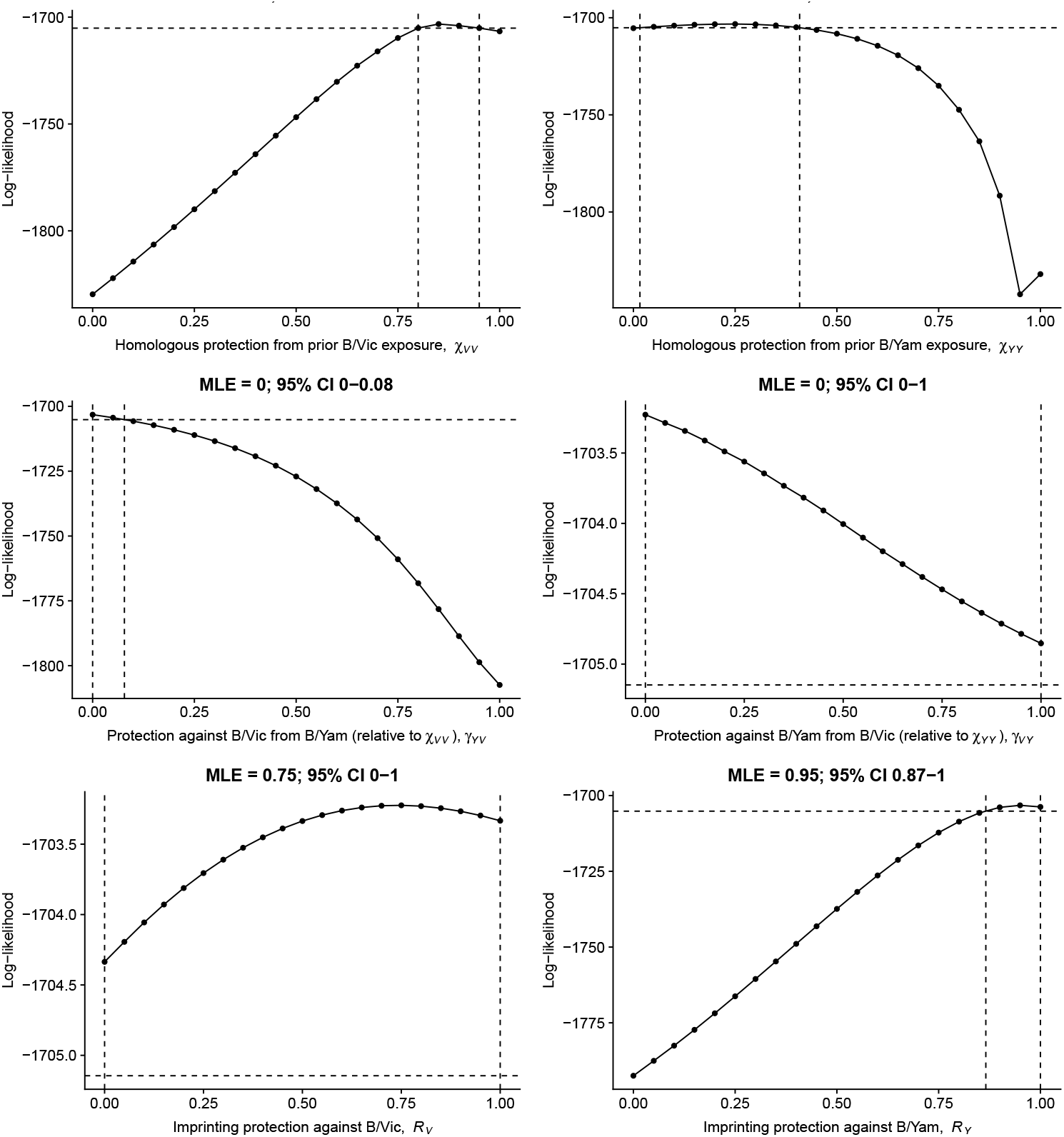
Likelihood profiles for protection parameters estimated from birth years starting in 1966 instead of 1959. The 95% confidence interval based on a likelihood ratio test with one degree of freedom is indicated by the vertical dashed lines.

**Figure S20.**
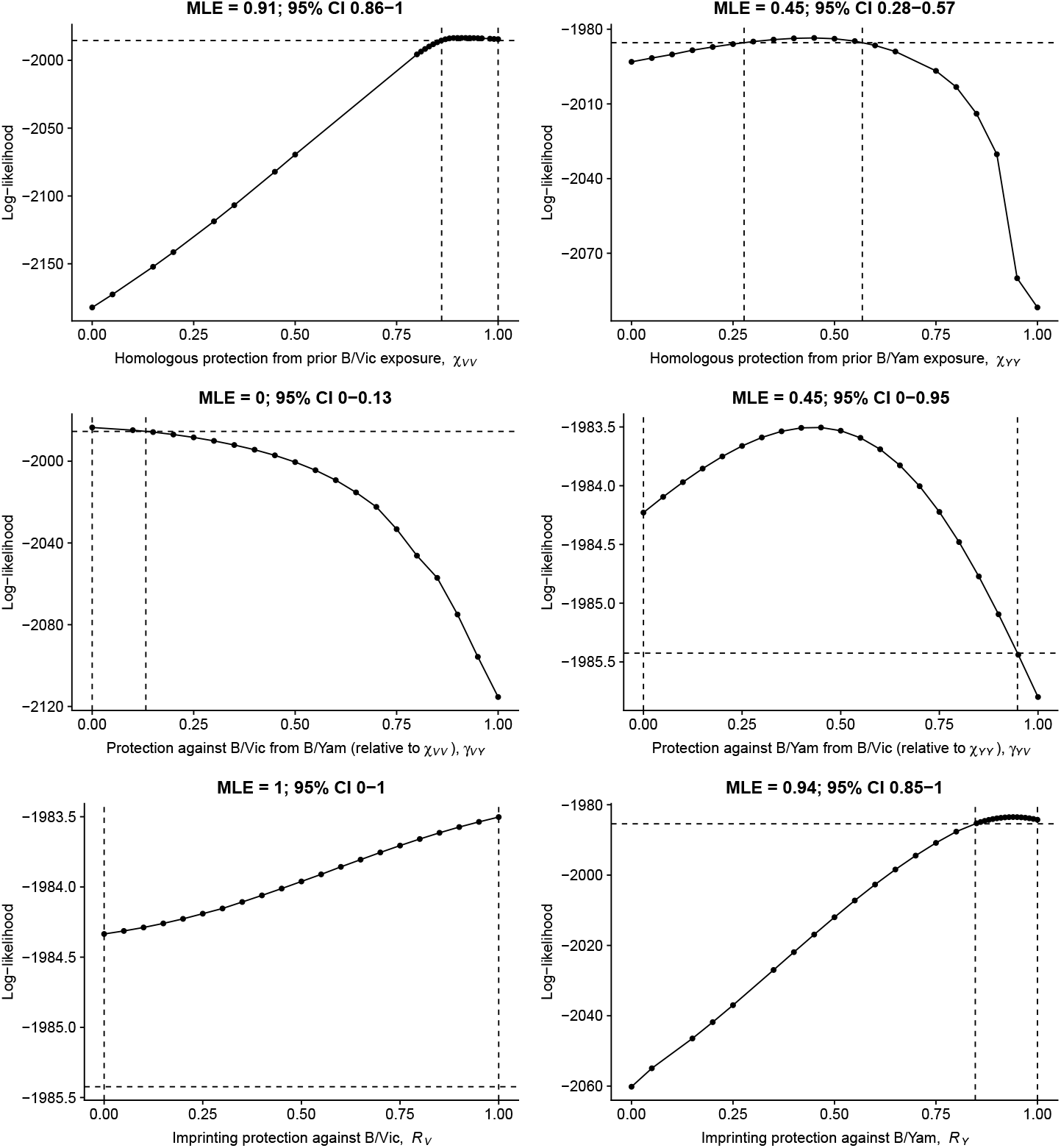
Likelihood profiles for protection parameters estimated by the model assuming a separate probability of medical attendance for infections in children under 5 years old. The 95% confidence interval based on a likelihood ratio test with one degree of freedom is indicated by the vertical dashed lines.

**Figure S21.**
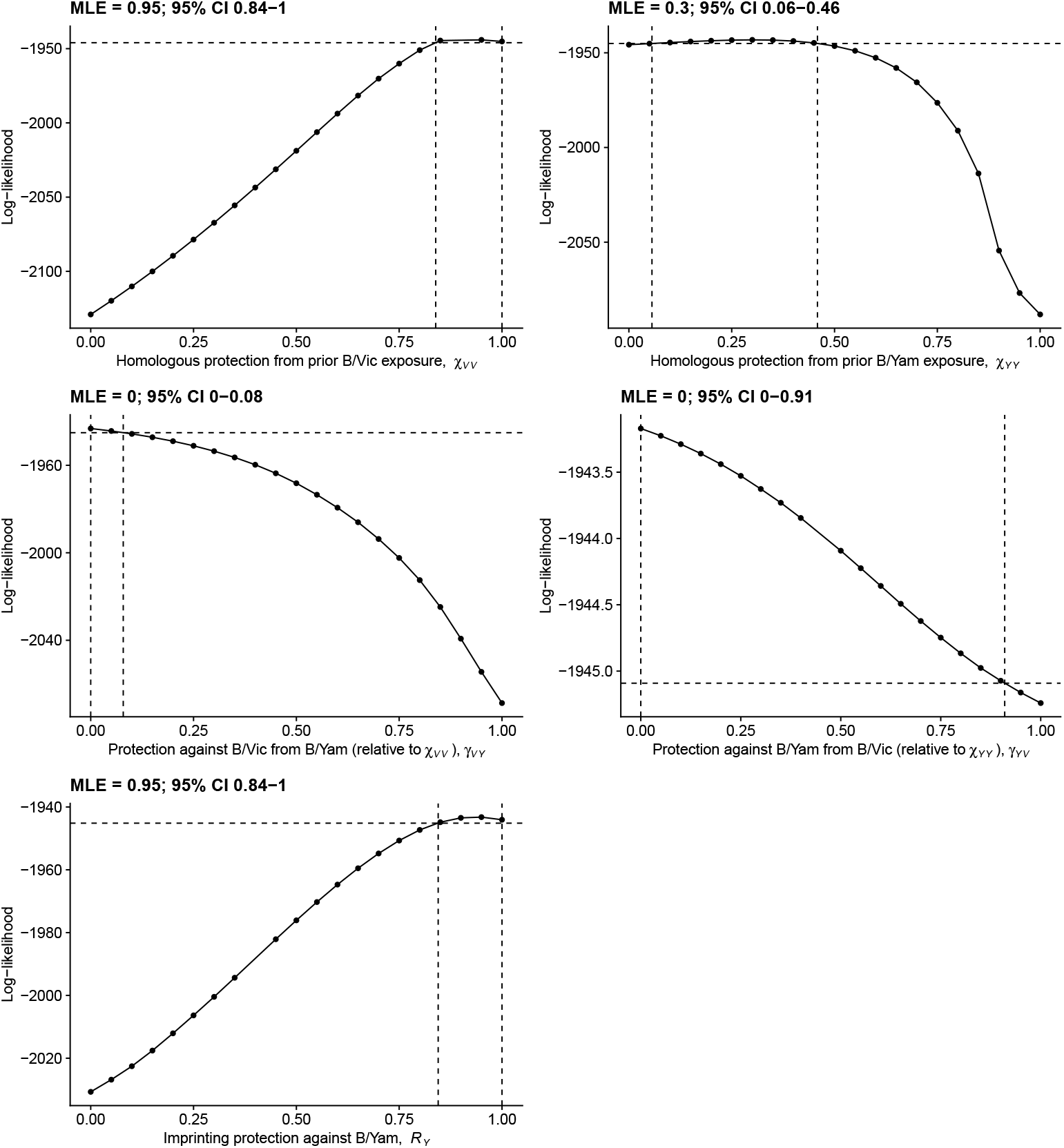
Likelihood profiles for protection parameters estimated from the complete New Zealand data after removing imprinting protection against B/Victoria and protection from strains circulating before 1988 against B/Yamagata. The 95% confidence interval based on a likelihood ratio test with one degree of freedom is indicated by the vertical dashed lines.

**Figure S22.**
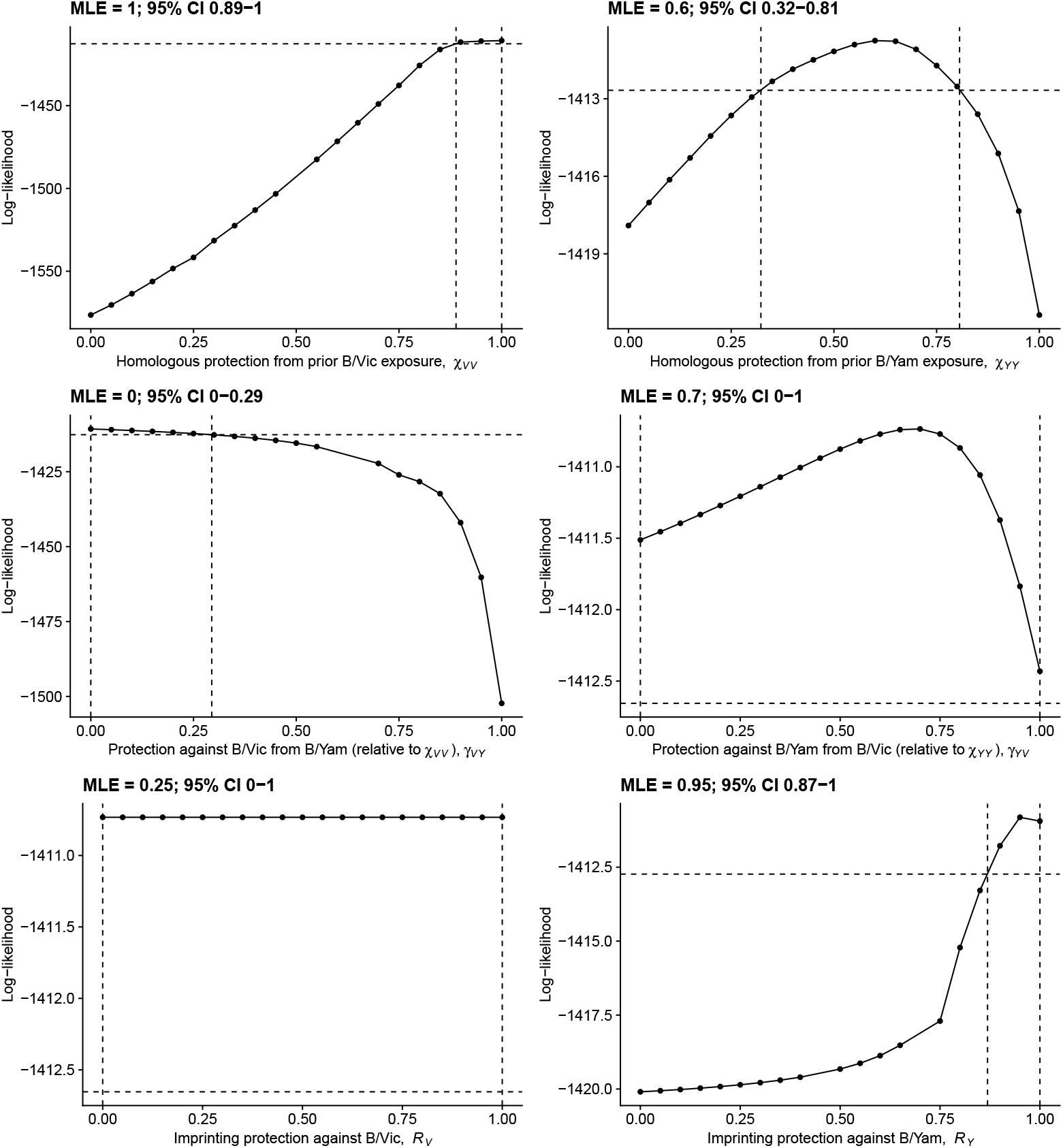
Likelihood profiles for protection parameters estimated after excluding children under 10 years old. The 95% confidence interval based on a likelihood ratio test with one degree of freedom is indicated by the vertical dashed lines.

**Figure S23.**
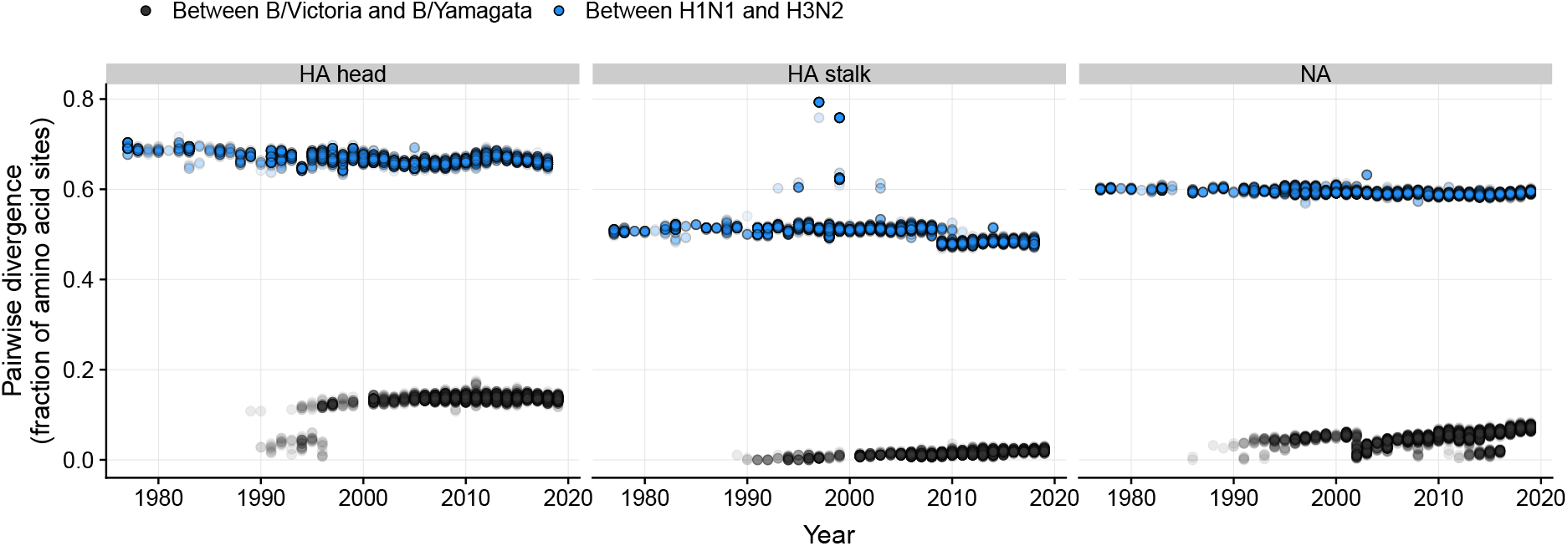
Amino acid divergence in the hemagglutinin and neuraminidase surface proteins between influenza A subtypes and between influenza B lineages. Divergence in hemagglutinin is shown separately for the antigenically variable head region (left) and the more conserved stalk region (center). Each point represents a pair of strains isolated in the same year and deposited on GISAID. When more than 100 isolates from a subtype or lineage were isolated in a single year, we used a random sample of 100 isolates, resulting in up to 4,950 pairs of each kind in each year. Up to 500 randomly chosen pairs are shown for each year. Average divergence values reported in the main text are based on the full set of pairs given the randomly sampled sequences.

**Figure S24.**
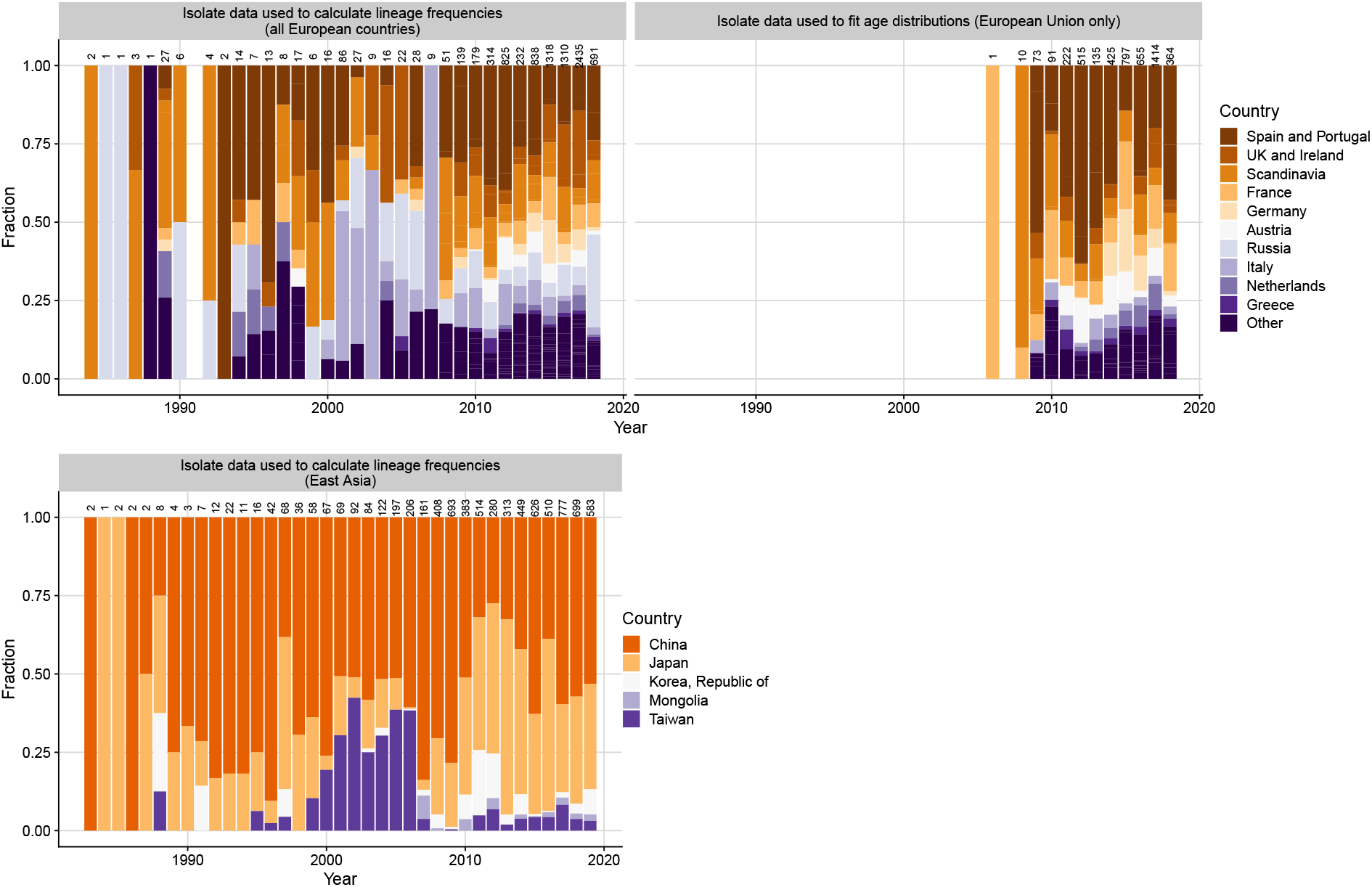
Geographic distribution of isolate data used to model the age distributions of influenza B cases in the European Union and China. We fitted the model to the age distribution of GISAID isolates in China and the European Union (EU). For those fits, historical lineage frequencies are also calculated from isolate data, including isolates that lack host age information. To increase statistical power, we used historical lineage frequencies estimated from all European countries represented on GISAID (including those not in the EU) when fitting to the EU age distributions (top left panel), and from all represented countries in East Asia when fitting to the Chinese age distributions (bottom left panel). The total numbers of isolates in each year are shown as numbers above the bars.

## Notes

### Competing Interest Statement

The authors have declared no competing interest.

### Author Declarations

Not applicable. Only aggregated case data previously collected by epidemiological surveillance were used in the analyses.

